# Health Impact Evaluation of Aspirational Districts Program in India: Evidence from National Family Health Survey

**DOI:** 10.1101/2023.07.27.23293263

**Authors:** Sandip K. Agarwal, Shubham Mishra

## Abstract

The Aspirational District Program (ADP) is a unique initiative of Government of India launched in 2018 that aims to reduce inter-district multidimensional inequality. ADP aims to bring the most backward districts to catch up with the rest of the other districts in the country. The program is comprehensive in its scope as it targets the improvement of several key development indicators spanning health and nutrition, education, agriculture and water resources, financial inclusion and skill development and basic infrastructure indicators. Aspirational districts (ADs) are eligible for enhanced funding and priority allocation of various initiatives undertaken by the central and state governments. Our research estimates the causal impact of ADP on the targeted health and nutrition indicators using a combination of propensity score matching and difference-in-differences (PSM-DID). We use the fourth and fifth rounds of National Family Health Survey (NFHS) data collected in 2015-16 and 2019-21 respectively which serve as the pre and post-treatment data for our analysis. Moreover, we take advantage of the transparent mechanism outlined for the identification of ADs under ADP, which we use for propensity score matching for our PSM-DID. While we observe negative impact of ADP on early initiation of breastfeeding, we believe that the impact is confounded with the effects of Covid-19 since part of NFHS-5 data was collected during the pandemic. Therefore, the negative impact of ADP on early initiation of breastfeeding disappears when we only use pre-covid data. Additionally, using pre-covid data we find a reduction in the prevalence of underweight children younger than 5 years by 2 to 4% in ADs as an impact of ADP, which is robust across multiple specification. We do not find evidence of any other positive or negative impact of ADP on any other health and nutrition indicators, which is robust. Future research efforts should be made toward impact evaluation of all the targeted indicators in order to get a comprehensive unbiased evaluation of ADP.

## 1 Introduction

Health inequality has been documented across countries as well as within a country across socioeconomic and disadvantaged groups as well as sub-national spatial units. In India and other low and middle-income countries (LMICs), while there have been several targeted interventions through social welfare and poverty alleviation schemes, the primary focus of such schemes has been to address deprivations, and therefore any implications of the scheme for addressing inequality are inadvertent. In 2018, the Government of India launched the ‘Transformation of Aspirational Districts’ or Aspirational Districts Program (ADP henceforth)^1^ with the objective of reducing regional multidimensional inequalities. ADP identified the most backward districts in the country and developed a comprehensive framework to prioritise development in these districts so that they can catch up with other districts in the country. While health inequality is not the only targeted dimension in ADP, health and education sectors equally contribute to 60% of the total outcomes. Other dimensions include agriculture and water resources, skill development, financial inclusion and basic infrastructure. Our research evaluates the impact of ADP on targeted health indicators in aspirational districts (ADs) before and after the policy intervention using the fourth and fifth rounds of the National Family and Health Survey (NFHS) data.

Experts have acknowledged that reproductive health, child health and nutrition are some of the greatest challenges for India^2^. Strong evidence of district-level inequality in maternal mortality rates (MMR) exists where MMR is measured as number of deaths per 100,000 live births. MMR ranges from 0 to 1671 across districts with a national average of around 142^3^. Similar evidence of high inequality in maternal health care utilization too exist among districts that includes indicators like antenatal and post natal care, institutional birth or delivery in the presence of skilled birth attendants^2,4–10^. While the national average for institutional births denoting percentage of children born in a health facility is 79%, the minimum across the districts is even less than 10%^11,12^. Evidence of huge geographical disparity in under-five child mortality rates (U5MR) and neo-natal mortality (NMR) rates also exist^13,14^. At the district level, U5MR varied 10·5 times between districts of India in 2017, ranging from 8·4 to 87·9 per 1000 livebirths with a national average of 42.4. Similarly, the neo-natal mortality rate (NMR) varied 8 times between the districts in India, with a range of 5·8 to 46·2 per 1000 livebirths and a national average of 23.5^14^. With regards to child immunization, while significant achievements have been made, full immunization is yet a distant thing, with several regions performing dismally low on vaccine preventable diseases^5,9,11,15–18^. The national average for percentage of children fully immunized is 63% but it is as low as 7% in the worst performing district. Child malnutrition too exhibits stark inequality across districts manifested in indicators measuring prevalence of stunting, wasting or underweight among children under five years of age^5,12,15–17,19^. Data from the fourth round of National Family Health Survey (NFHS-4) collected in 2015-16 indicates that percentage of stunted, wasted and under-weight children in a district is as high as 65%, 67% and 47% respectively quite disparate from the national averages of 36%, 21% and 33% respectively. Recent research has drawn attention to heterogeneity in health and nutrition indicators using micro-level data at sub-district level even at the level of villages based on which researchers have argued for geographical targeting at micro-spatial units^20–22^.

In the context of developing countries, particularly India, although there has been an increase in efforts toward research on health inequity in the past two decades, research studies have predominantly focused on documenting health disparities^23,24^. While it is necessary to know the status and trends of health disparities in the country, it is not sufficient to design health policies that would reduce health disparities. Instead information on health disparities would rather help policymakers in identifying the health indicators and the regions that needs to be targeted rather than providing solutions to address health inequities. Impact evaluation of health policies have exhibited substantial heterogeneity in causal impact of health policies across different subgroups and sub-national spatial units. For example, impact evaluation of Janani Suraksha Yojana (JSY), a conditional cash transfer scheme in India for promoting institutional births has found substantial inter-state and inter-district variation in the impact of the program on institutional deliveries, skilled birth attendance, breast feeding behavior and child immunization rates^25–29^. The heterogeneity in the impact of JSY across districts is not limited to differences in magnitude of the impact but also stands in stark contrast in terms of direction of the impact across districts. New evidence has surfaced that while the average impact of JSY on perinatal mortality is zero, differences in outcome exist between districts that operate at below-median preexisting capacity in the secondary health care system and districts with above-median capacity with JSY leading to an increase in the risk of perinatal mortality in the former^30^. In another impact evaluation of a statewide maternal and child health program in Bihar in India, Ward et al. (2020)^31^ found the program to have a variable impact on the existing inequity between the least and the most marginalised groups across the continuum of care and delivery platforms. The program did not narrow the health inequity across most health-related behaviours, instead there were significant increase in inequity across health indicators that relied upon access to health care. Researchers have emphasized on the value of impact evaluation of health policies at sub-national units and for sub-groups in order to understand the underlying reasons for heterogeneous impact of health policies which would be useful for formulating effective policy guidelines^26,32,33^. Health programs should be equity sensitive by not just simply measuring the underlying disparities but also targeting them by monitoring progress throughout implementation^31^.

Our research makes an important contribution by assessing the impact of a policy exclusively designed to promote inclusive development by reducing multidimensional regional inequality. In order to ensure that large-scale health interventions are closing gaps of disparity and promote inclusive health advancements, governments must be willing to invest in equity oriented health systems^31^. ADP by the Government of India are conscious efforts made in this direction. We evaluate ADP in India from the lens of health inequity and discuss our findings, which we believe would be of interest to researchers, practitioners and policy makers. Using NFHS-4 data, Subramanian et al.^34^ developed a metric that takes into account prevalence and (population weighted) headcount of various child health and anthropometric measures, which was used to identify the worst performing districts for prioritization of interventions for child health and nutrition. Several of these districts also have some degree of overlap with the ADs making it pertinent to evaluate the health outcomes in ADs. To the best of our knowledge, our research is the first to find the causal impact of ADP. In an earlier policy paper by Green and Kapoor^35^, although authors attempt to assess the ADP by accounting for changes in targeted indicators, the analysis is not causal, which the authors acknowledge. Similarly, in another research paper, Subramanian et al. (2023)^36^ evaluated Indian districts including the ADs using data from NFHS-4 and NFHS-5, where the assessment was to predict the progress of a district in terms of achieving 33 Sustainable Development Goal (SDG) indicators by 2030. Similarly, Let et al. (2024)^37^ assess the prevalence of anaemia among women of reproductive age in ADs using data from fourth and fifth rounds of NFHS. However, none of the above research establishes casual evidence of the impact of ADP. We fill this gap by estimating the causal impact of ADP on targeted health indicators. Our paper is structured as below. Section 2 provides a brief description of the ADP followed by data and the methodology in section 3. Results of our analysis are presented in section 4 followed by a discussion of the findings in section 5 and conclusion in section 6 respectively.

## 2 Aspirational Districts Program (ADP)

The ‘Transformation of Aspirational Districts’ or Aspirational Districts Program (ADP) was launched by the Government of India in January 2018 with the objective of achieving inclusive development through a reduction in multidimensional inequalities by focusing on districts that have been lagging in several development indicators. Although, ADP is comprehensive and not limited to health indicators, health and nutrition is a key sector for the identification of backward districts as well as targeted program outcomes. The targeted outcomes are 49 performance indicators that include 81 data points, of which 13 indicators covering 31 data points are from the health domain itself. The remaining indicators measure performance related to education, agriculture and water resources, financial inclusion and skill development and basic infrastructure.^1^

Aspirational districts (ADs) are eligible for enhanced funding and priority allocation of various initiatives undertaken by the central and the state governments. Three key mechanisms underlying ADP that are expected to drive ADP toward its goal are convergence (of central and state schemes), collaboration (of central, state-level officers and district collectors) and competition among districts driven by a mass movement. Researchers have strongly argued for purposeful investment by policymakers in equitable health programs by measuring, addressing and targeting the underlying disparities with continuous monitoring of progress throughout implementation^31^. A robust continuous monitoring and evaluation framework is a key feature of ADP towards building of inclusive health policies, which has been a major shortcoming of past initiatives^38^.

ADP is a multi-sector program where sector specific operational guidelines have been devised by the respective departments to meet the objectives of ADP. In the context of the health sector, emphasis has been made on intensifying the actions in ADs through a continuous process of situational analysis, health plan, implementation of key programs and finally monitoring and sustenance for the respective AD^39^. Situational analysis identifies the gap in the health services at the lowest level which includes accessibility of health services, quality of health services through means of verification, availability of essential medicines and health workforce and utilization of sanctioned funds for the AD. Based on the outcomes of the situational analysis, a district level health plan is developed to achieve the low hanging fruits in terms of health outcomes and also tailor the interventions as per the local context making room for participation from various stakeholders. Additionally, ADs are provided with the flexibility to use non-health district funds like district mineral fund, district tribal fund, minorities development fund etc. for intensification of health activities at the lowest level. The operational guidelines builds a robust mapping of different interventions under key programs to the health indicators targeted under ADP for potential improvement^391^. Lastly, the targeted outcomes are rigorously and continuously monitored and reviewed at the lowest level in order to sustain the impact of interventions in ADs.

ADs have been identified through a transparent mechanism based on the ranking of a composite index developed from 11 core measurable indicators summarized in the table 1. Health indicators have been assigned one of the highest weights (contributing to 30%) in the composite index that includes four variables - antenatal care, institutional deliveries, stunting and wasting of children below five years where each of them have equal weights. The other sectors used for construction of the composite index include infrastructure with 30% weightage, deprivation measured by landless household dependent on manual labor contributing one-fourth to the index, and the remaining 15% is contributed by education. As it can be seen in table 1,

**Table 1.** Variables used to identify ADs (Source: Aspirational Districts Primer^1,39^.

| Indicator | Source | Sector | Weight |
| --- | --- | --- | --- |
| Landless household dependent on Manual Labour | SECC D7 | Deprivation | 25% |
| Ante Natal Care | NFHS-IV | Health | 7.5% |
| Institutional Deliveries | NFHS-IV | Health | 7.5% |
| Stunting of children below 5 years | NFHS-IV | Health | 7.5% |
| Wasting of children below 5 years | NFHS-IV | Health | 7.5% |
| Elementary Drop-out Rate | U-DISE 2015-16 | Education | 7.5% |
| Adverse pupil teacher ratio | U-DISE 2015-16 | Education | 7.5% |
| Unelectrified household | Ministry data | Infrastructure | 7.5% |
| Household without individual toilet | Ministry data | Infrastructure | 7.5% |
| Un-connected PMGSY village | Ministry data | Infrastructure | 7.5% |
| Rural Household without access to water | Ministry data | Infrastructure | 7.5% |

ADP guidelines also mention the data source used for identifying the ADs along with the indicators (measured in percentages unless explicitly stated)^1,39^. The data for health variables are taken from the fourth round of NFHS, deprivation from the Socio Economic Caste Census (SECC) data, education from Unified District Information System on Education (UDISE) and infrastructure variables from the respective ministries. Using the above mechanism, the composite index for every district is constructed and districts are ranked based on this composite index to arrive at 117 most backward districts such that each state has at least 1 district as part of the ADP. West Bengal declined to be part of the ADP, hence after excluding 5 districts of West Bengal, there are 112 ADs^35^. Figure 1 maps the AD in comparison to the non-ADs on the map of India.

**Figure 1.**
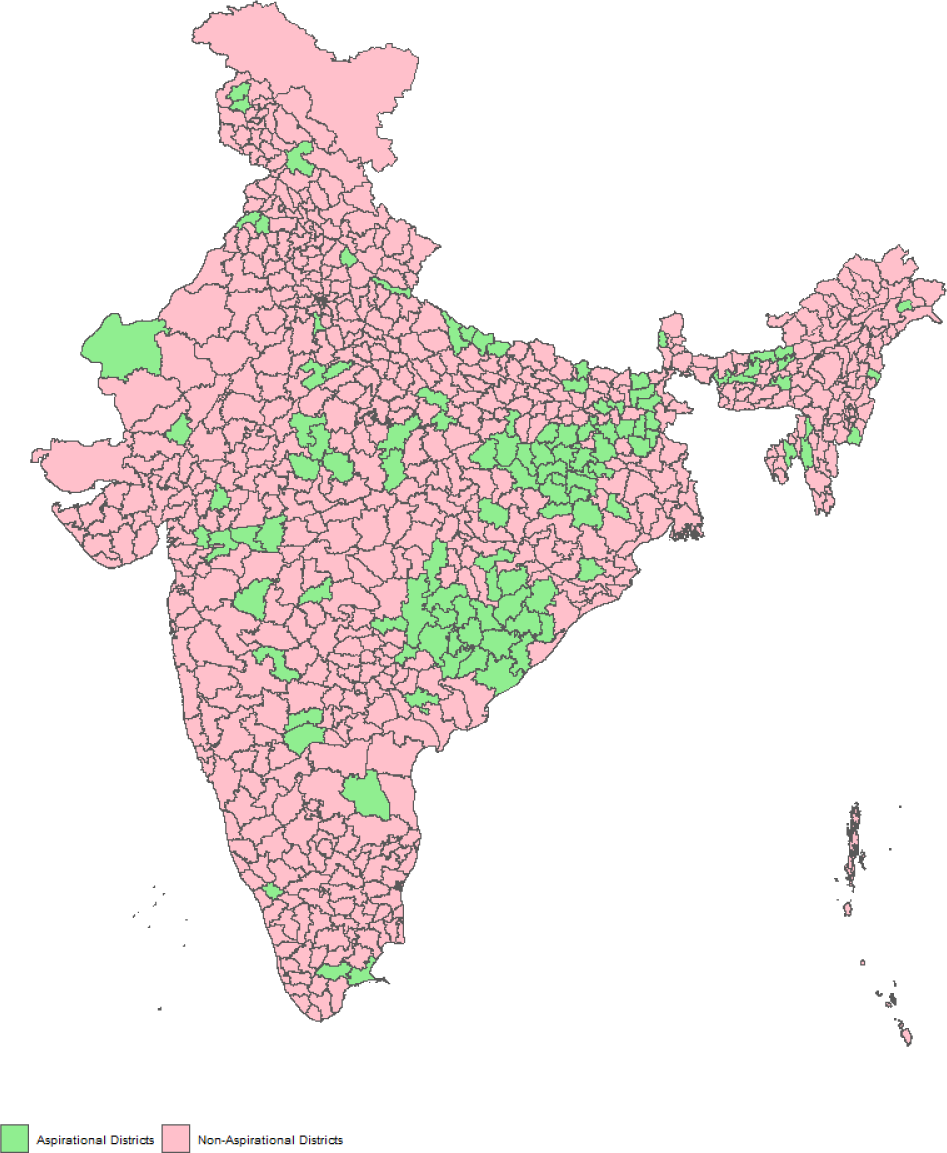
Aspirational Districts in India

A key feature of ADP is continuous monitoring and evaluation of targeted indicators. The performance metrics contribute to the overall and sector-specific ranking of a district, which is publicly available on the website Champions of Change dashboard of ADP (http://championsofchange.gov.in/site/coc-home/). This is expected to foster healthy competition among the ADs, which drives their monthly rankings. Several ADP targeted indicators are collected every few years that inhibits continuous monitoring. Therefore, such indicators are measured through survey or self-reported values of districts (non-validated or survey validated). While, the above approach is indispensable for continuous monitoring, yet relying solely upon them for assessing the impact of ADP could lead to a biased evaluation. Self reported indicators by districts are often not validated, which could be upward biased due to the competition faced by the districts. Moreover, causal impact evaluation of ADP, require measurement of outcomes for non-ADs too. Therefore, a nationally representative health outcome data is required for estimating the casual impact of ADP on health indicators.

We use district-level NFHS data to estimate the impact of ADP on targeted health outcomes. NFHS data have been a trusted and reliable source of health data for decades among researchers, practitioners and policymakers, which has shaped important health policy decisions for the country. NFHS data collection is not motivated to reflect any desirable outcomes of ADP but rather centered around the objective of tracking the health status of the population in the country. Therefore, using NFHS data for the health impact evaluation of ADP is unlikely to suffer from desirability bias unlike the self-reported district performance indicators. Secondly, there are no large methodological differences in execution of NFHS data collection that would make inter-district comparison inconsequential. Finally, NFHS is nationally representative, and we are able to obtain estimates of health indicators for all districts irrespective of whether AD status was assigned to a district or not. Therefore, access to the fifth round of NFHS data facilitated the health impact evaluation of ADP.

## 3 Data and Methods

We estimate the causal impact of ADP on targeted health and nutrition outcomes by employing fourth and fifth rounds of district-level NFHS data. We take advantage of the timing of the launch of ADP in 2018, which was between NFHS-4 and NFHS-5 data that allows us to use the fourth and the fifth rounds of NFHS data as the pre and post-intervention data respectively for comparing the changes in ADP-targeted health outcomes.

We use a combination of propensity score matching (PSM) and difference-in-differences (DID) to estimate the causal impact of ADP on targeted health and nutrition outcomes. A growing body of literature in health economics and policy evaluation has employed the combination of PSM and DID (PSM-DID) to estimate the causal impact of a policy intervention using observational data^40–44^. A reasonable choice of model for impact evaluation of ADP would have been a DID model given the availability of NFHS-4 and NFHS-5 data that could serve as pre- and post-intervention data and ADs and non-ADs as treatment and control units. We know that validity of DID estimates rests crucially upon the parallel trend assumption implying that the pre-treatment trend of the outcome variable between the control and the treatment group are parallel. In order to test for the parallel trend assumption more than one period of pre-treatment data is needed. However, we are unable to test for pre-treatment parallel trends due to data limitations, as NFHS data are not available for districts but only for the states prior to NFHS-4^45^. Therefore, to circumvent this problem, we use PSM before implementing the DID so that we are able to create valid counterfactual for our treatment districts before.

PSM has been widely used for causal impact evaluation in observational studies^28,46–49^. In PSM, the probability of treatment assignment depends on observed covariates, which are used to estimate the propensity scores (PS). The estimated PS is matched between the treatment and the control units to create a valid counterfactual group for the treatment units. PSM enables the design and analysis of observational studies to mimic certain aspects of randomized controlled trials. Essentially, the PS acts as a balancing score by ensuring that the distribution of observed baseline covariates is similar between the treated and untreated units given their PS values^49^. It allows us to estimate the Average Treatment Effect in Treated (ATT)^50^ for an intervention. Using PSM before DID enables comparison between the groups, and strengthens the plausibility of the parallel trends assumption, thus reducing the selection bias and dependence upon the unobservables and observables consistent over time or having similar trends between the treatment and the counterfactual^40,44^. In line with Heckman, Ichimura, and Todd (1997)^51^, PSM-DID method allows for estimation of temporally invariant unobserved outcome differences between individuals in the treatment and control groups, akin to fixed effects in panel data analysis. Therefore, we use the PSM-DID method for impact evaluation of the ADP on potentially targeted health and nutrition outcomes present in NFHS data.

### 3.1 PSM-DID

We would like to estimate the average treatment effect on the treated (ATT) on selected outcome variables, which would provide us with the casual estimate of the impact of ADP. Following Cheng et al.(2015)^52^, the ATT is given by

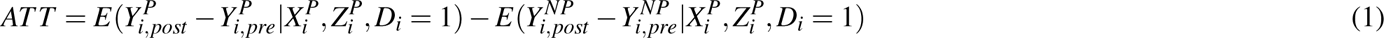

where 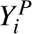 and 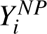 denotes the treated and the non-treated outcomes for district *i* with an additional subscript *pre* and *post* indicating time period for pre or post intervention. The outcomes are conditional upon a set of observed and unobserved attributes of the district denoted by *X_i_* and *Z_i_* respectively. *D_i_* is an indicator variable with value 1 for treatment else 0. While, 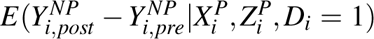 is unobserved, a fundamental assumption in the matching literature is that the treatment assignment can be assumed to be random if the treatment and the control groups are matched on the observed covariates, i.e. *X^P^* = *X^NP^* = *X*, which implies that 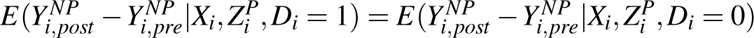. Therefore, the ATT can be rewritten as

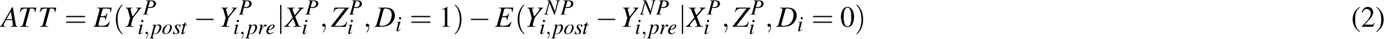

In the above equation, we can also get rid of the unobservables, if we believe that *Z_i_* are time invariant or have the same time trend between the treated and the untreated. Moreover, given the assumption of conditional Independence, the potential outcomes are independent of the treatment status when matched on the covariates. Therefore, the ATT can be estimated conditional on the propensity scores instead of conditional upon observed covariates.

The assumption of conditional independence also known as strict exogeneity or unconfoundedness assumption or ignorable treatment assignment^28,50,53^ is a fundamental assumption that must hold for the PSM estimates to be valid. The strict exogeniety assumption states that potential outcomes are orthogonal to treatment status conditional upon the observable covariates. In other terms, conditional on observable characteristics, participation is independent of potential outcomes. While, our PSM-DID analysis uses data for two time periods - one before the intervention and the other post intervention, we use only the pre-intervention data for implementing the PSM method followed by DID. However, the variables used for estimation of propensity scores and matching could not be used as an outcome variable as it is likely to violate the strict exogneity assumption that might result in biased estimates. Therefore, we excluded indicators that were used for program assignment at the baseline for impact evaluation of ADP even if they have been one of the targeted health indicators of the ADP. Hence, our PSM-DID implementation satisfies the strict exogneity assumption.

PS or the probability of treatment assignment is estimated using a logit regression as in equation 3 where *D_i_* is the binary variable with value 1 if the *i^th^*district is an AD else 0. *X* is the vector of observed covariates.

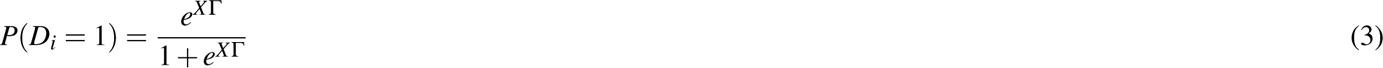

In the context of ADP, we have information regarding the treatment assignment criteria. Therefore, we know the set of *X* s or the observed covariates that were employed for treatment assignment as described in table 1. We use the same for matching on covariates in order to find a close counterfactual for every treated district. Subsequently, the ATT conditional upon PS can instead be written as

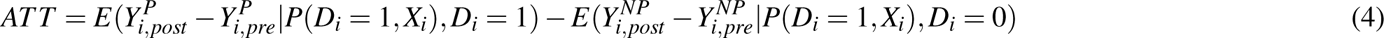

We use full matching method for matching the PS to find a counterfactual for every treated unit. In this method, each treated unit is matched with one or more untreated units based on their PS by minimizing the matched sample’s total absolute within-subclass distances by the number of sub-classes selected and the units assigned to each subclass.^49,54^. Full matching method has been demonstrated to be an effective matching technique for reducing bias due to observed confounding variables^48^.

### 3.2 Evaluation Outcomes

The targeted health and nutrition outcomes of ADP consists of 13 core indicators along with 31 sub-indicators that are listed in the ADP guidelines^1,39^. The full list of 31 health indicators targeted for improvement under ADP are provided in table S1 in the appendix. We restrict our analysis to those health indicators that have a corresponding variable measured in NFHS data. Out of 13 core health indicators, 3 indicators related to anaemia among pregnant women, tuberculosis and health infrastructure are not available in NFHS, and thus could not be utilized in our analysis. Among the remaining 18 sub-indicators (across 10 core indicators), we further exclude indicators that have been used for program assignment based on pre-intervention data from NFHS-4 so in order to satisfy the strict exogneity assumption of the PSM-DID. For one of the indicators on full immunization of children, NFHS-5 recorded two variables one based on information from vaccination card and the other based both vaccination card and mother’s recall. We included both the indicators in our analysis ^2^. Table 2 summarizes the mean of health indicators for AD and non-ADs pre and post intervention (i.e. for NFHS-4 and NFHS-5) along with p-values stating the statistical significance for the difference of means between AD and non-AD.

**Table 2:** Summary Statistics of ADP targeted health indicators pre and post intervention.

| No. Indicators | NFHS-5 |  |  | NFHS-4 |  |  |
| --- | --- | --- | --- | --- | --- | --- |
|  | Non-AD Mean<br>(n=516) | AD Mean<br>(n=105) | p-val. | Non-AD Mean<br>(n=516) | AD Mean<br>(n=105) | p-val. |
| 1.3 Registered pregnancies for which the mother received a Mother and Child Protection (MCP) card | 96.34 | 95.55 | 0.10 | 89.29 | 87.80 | 0.18 |
| 2 Mothers who consumed iron folic acid for 100 days or more when they were pregnant | 45.87 | 40.40 | 0.01 | 31.31 | 23.83 | 0.00 |
| 4.1 Sex ratio at birth for children born in the last five years | 948 | 943 | 0.74 | 932 | 943 | 0.32 |
| 5 Home births that were conducted by skilled health personnel | 2.76 | 5.17 | 0.00 | 3.71 | 5.77 | 0.00 |
| 6.1 Children under age 3 years breastfed within one hour of birth | 45.20 | 40.91 | 0.03 | 44.96 | 45.36 | 0.82 |
| 7 Children under 5 years who are underweight (weight for age) | 28.44 | 35.92 | 0.00 | 31.53 | 40.93 | 0.00 |
| 8.2 Children with diarrhoea in the 2 weeks preceding the survey who received oral rehydration salts (ORS) | 17.77 | 24.34 | 0.05 | 53.42 | 53.27 | 0.95 |
| 8.3 Children with diarrhoea in the 2 weeks preceding the survey who received zinc | 9.13 | 12.52 | 0.07 | 22.19 | 20.57 | 0.30 |
| 8.4 Children with fever or symptoms of ARI in the 2 weeks preceding the survey taken to a health facility or health provide | 43.60 | 48.35 | 0.15 | 72.04 | 69.47 | 0.10 |
| 10.1 Breastfeeding children age 6-23 months receiving an adequate diet | 11.77 | 11.14 | 0.41 | 9.72 | 8.80 | 0.20 |
| 10.2 Non-breastfeeding children age 6-23 months receiving an adequate diet | 1.18 | 1.12 | 0.89 | 14.89 | 14.89 | 1.00 |
| 11 Children age 12-23 months fully vaccinated based on information from either vaccination card or mother's recall | 76.74 | 76.80 | 0.96 | 62.61 | 59.53 | 0.10 |
| 11 Children age 12-23 months fully vaccinated based on information from vaccination card only | 81.85 | 81.78 | 0.96 | 62.61 | 59.53 | 0.10 |

## 4 Results

### 4.1 PSM

In observational studies using PSM researchers often do not have the program assignment rule. Therefore, PSM is implemented by using a list of covariates for matching that reduces the bias between the treated and the comparison group. In the case of ADP, program assignment for the districts to be included as ADs is based on a composite index developed from a set of covariates described in table 1. We take advantage of this transparent mechanism by using the same variables for matching that were used for program assignment. We use pre-intervention data for all variables in Table 1 for PS estimation for treatment assignment. Although we used data from sources listed in Table 1 for most of the variables, for a few variables that used ministry data we used indicators from alternative sources as the ministry data was not available. ‘Household without individual toilet’ and ‘Rural Household Without Access to Water’ was captured by the inverses of ‘Percent of Population living in households that use an improved sanitation facility’ and ‘Percent of Population living in households with an improved drinking water source’ from NFHS-4 data source respectively. Few variables used for program assignment have multiple indicators in NFHS-4, which measure different intensities of the same variable. For example, NFHS-4 has three variables for antenatal care, of which the first one is the percentage of mothers who had full antenatal care, the second measures one antenatal checkup in the first trimester and the third measures atleast four antenatal care visits. Similarly, we also have two indicators for children wasted - percentage of children wasted and percentage of children severely wasted. We use all these variables for estimating PS. For Unconnected PMGSY village, we use data on number and length of roads planned under PMGSY (Pradhan Mantri Gram Sadak Yojana), which is the rural roads programme in India implemented in three phases - PMGSY-I (2000), PMGSY-II (2013) and PMGSY-III (2019) ^3^. PMGSY-I and II are pre-intervention and PMGSY-III is post-intervention. We use additional roads sanctioned in 2019 under PMGSY-III as an indicator of unconnected PMGSY villages before the ADP intervention. As indicators of unconnected PMGSY villages, we use number of roads and length of road in kms sanctioned under PMGSY-III in 2019 as percentages over the total across all the three phases. All the above variables are used as predictors of program assignment in the logit regression for the estimation of PS. Subsequently, the estimated PS are matched using full matching in order to create a valid counterfactual for the treated districts.

We have a total of 599 districts for our analysis of which 104 are ADs. Table 3 provides the summary of balance between AD and non-ADs. We implement full matching on variables using pre-intervention data as described earlier. We are left with 92 ADs and 389 non-ADs after full matching based on estimated propensity scores, which satisfies the assumption of common support. The assumption of common support is a crucial assumption for making meaningful comparisons across control and treatment units. Table 4 summarizes the balance of statistics between ADs and non-ADs after matching. Figure 2 plots the density of observed covariates across ADs and non-ADs before and after matching. The scatterplot of estimated propensity scores for comparison between ADs and non-ADs is provided in figure S1 in the appendix.

**Figure 2.**
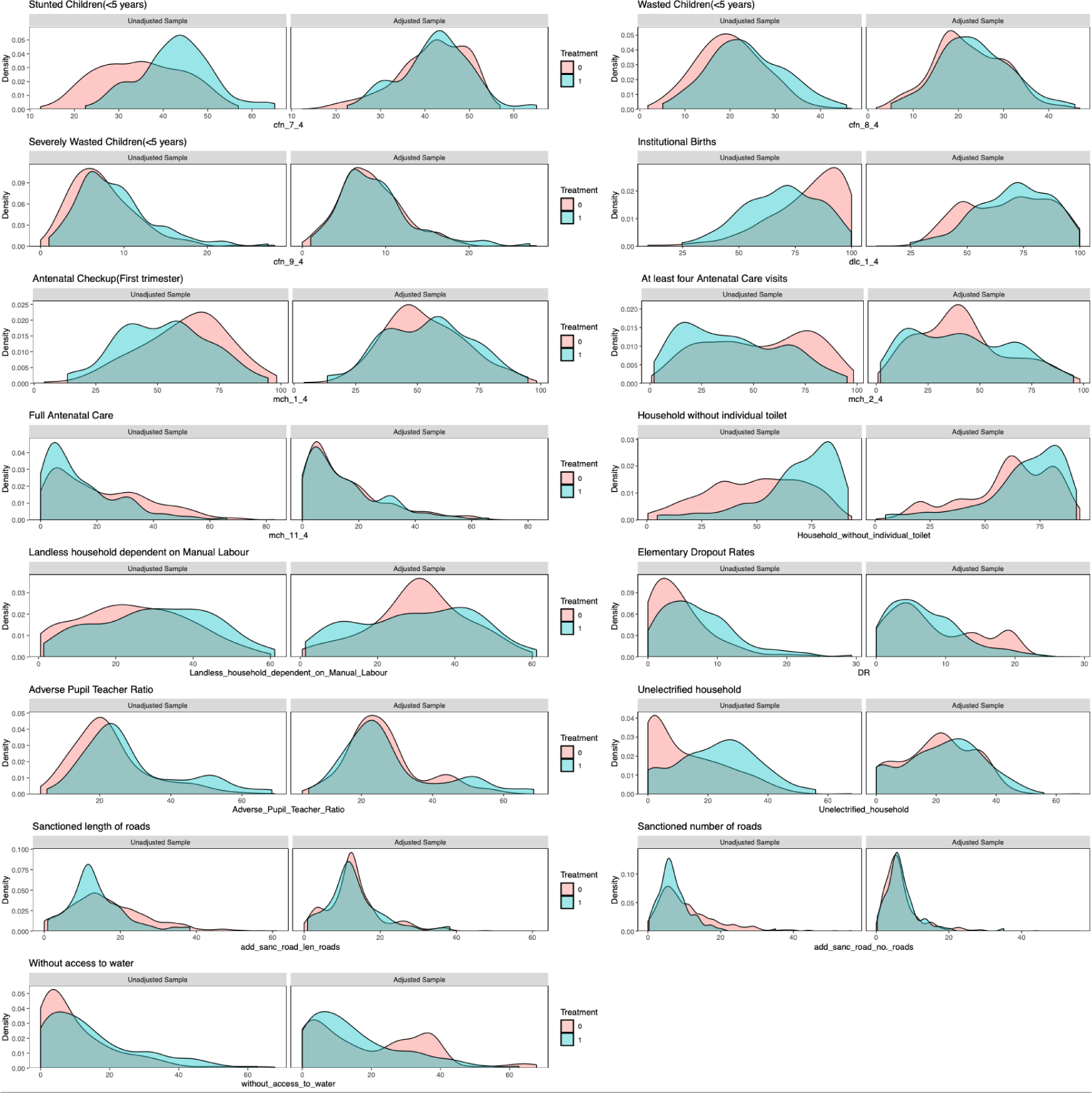
Density plots across AD and non-AD for co-variates before and after matching

**Table 3.** Summary of Covariate Balance before matching.

| Variable | Mean |  | St. Mean | Var. | eCDF |  |
| --- | --- | --- | --- | --- | --- | --- |
|  | AD<br>(n = 104) | Non-AD<br>(n = 495) | Diff. | Ratio | Mean | Max |
| Landless household dependent on Manual Labour | 30.2397 | 24.5434 | 0.3830 | 1.1236 | 0.1101 | 0.1911 |
| Antenatal checkup in the first trimester | 52.5760 | 60.9505 | -0.4650 | 1.0261 | 0.1256 | 0.2373 |
| At least 4 antenatal care visits | 39.9740 | 52.6638 | -0.5295 | 0.8942 | 0.1419 | 0.2277 |
| Full Antenatal Care | 14.6356 | 21.6954 | -0.5230 | 0.6458 | 0.1273 | 0.2088 |
| Institutional Births | 68.8885 | 80.3258 | -0.6842 | 0.9943 | 0.2040 | 0.3498 |
| Stunted Children(<5 years) | 42.8942 | 34.8777 | 0.9746 | 0.7249 | 0.2090 | 0.3858 |
| Wasted Children(<5 years) | 23.8192 | 20.1223 | 0.4390 | 1.2840 | 0.1105 | 0.1886 |
| Severly Wasted Children(<5 years) | 8.9558 | 7.4608 | 0.3199 | 1.3374 | 0.0788 | 0.1577 |
| Elementary Dropout rates | 7.4513 | 4.6893 | 0.4976 | 1.5501 | 0.1776 | 0.2982 |
| Adverse Pupil Teacher Ratio | 28.9527 | 22.8524 | 0.4356 | 1.6738 | 0.1402 | 0.2458 |
| Unelectrified household | 23.1067 | 12.8397 | 0.7634 | 1.1093 | 0.2287 | 0.3641 |
| Household without individual toilet | 67.9798 | 49.2710 | 0.9761 | 0.7684 | 0.2416 | 0.4272 |
| Household without access to water | 15.3510 | 10.9193 | 0.3178 | 1.3277 | 0.1068 | 0.2083 |
| PMGSY-III sanctioned number of roads | 7.3158 | 10.7133 | -0.7052 | 0.3301 | 0.1153 | 0.2441 |
| PMGSY-III sanctioned length of roads | 13.6389 | 16.8888 | -0.4423 | 0.4998 | 0.0989 | 0.1949 |

**Table 4.** Summary of Covariate Balance after matching.

| Variable | Mean |  | St. Mean | Var. | eCDF |  |
| --- | --- | --- | --- | --- | --- | --- |
|  | AD<br>(n = 92) | Non-AD<br>(n = 389) | Diff. | Ratio | Mean | Max |
| Landless household dependent on Manual Labour | 29.9880 | 30.8785 | -0.0599 | 1.5267 | 0.0734 | 0.1434 |
| Antenatal checkup in the first trimester | 54.2337 | 52.9844 | 0.0694 | 1.2113 | 0.0429 | 0.1224 |
| At least 4 antenatal care visits | 41.6065 | 41.3533 | 0.0106 | 1.2536 | 0.0471 | 0.1370 |
| Full Antenatal Care | 15.3326 | 15.5384 | -0.0152 | 0.9498 | 0.0218 | 0.0869 |
| Institutional Births | 70.7402 | 67.7892 | 0.1765 | 0.7787 | 0.0426 | 0.1871 |
| Stunted Children(<5 years) | 42.3326 | 41.6082 | 0.0881 | 0.9964 | 0.0246 | 0.0869 |
| Wasted Children(<5 years) | 23.7467 | 22.3381 | 0.1673 | 1.0822 | 0.0400 | 0.1569 |
| Severely Wasted Children(<5 years) | 8.9174 | 8.3997 | 0.1108 | 1.4062 | 0.0235 | 0.0797 |
| Elementary Dropout rates | 7.0187 | 8.4655 | -0.2606 | 0.6234 | 0.0464 | 0.1838 |
| Adverse Pupil Teacher Ratio | 28.0760 | 26.3755 | 0.1214 | 1.7462 | 0.0374 | 0.1149 |
| Unelectrified household | 22.1237 | 20.5387 | 0.1179 | 1.1629 | 0.0339 | 0.1462 |
| Household without individual toilet | 67.2652 | 60.7130 | 0.3418 | 0.9096 | 0.0923 | 0.2485 |
| Household without access to water | 15.4185 | 19.7731 | -0.3123 | 0.6627 | 0.0859 | 0.2197 |
| PMGSY-III Sanctioned number of roads | 6.9545 | 7.0009 | -0.0096 | 0.7882 | 0.0289 | 0.1281 |
| PMGSY-III Sanctioned length of roads | 13.1564 | 13.2235 | -0.0091 | 0.9310 | 0.0226 | 0.1536 |

### 4.2 PSM-DID

The estimates of ATT from PSM-DID analysis for the targeted health and nutrition outcomes (as summarized in table 2 are presented in table 5. We do not find a statistically significant impact of ADP on any of the targeted health and nutrition outcomes except for an adverse effect on children under age 3 years breastfed within one hour of birth (also known as early initiation of breastfeeding). The impact of ADP on early initiation of breastfeeding is negative with a reduction of around 6 percentage points statistically significant at 1% level of significance. It is possible that the above result is a matter of chance as we are testing multiple hypothesis. Therefore, we also report adjusted p-values following the Benjamini-Hochberg (BH) procedure in order to control for false discovery rate (FDR)^55^. The statistically significant negative impact of ADP on early initiation of breastfeeding does not survive FDR as evident from the adjusted p-values for multiple hypothesis testing following the BH procedure.

**Table 5.** Results of PSM-DID.

| No. | Indicators | Est. | St. err. | t-stat. | p-val. | Adj.<br>p-val. |
| --- | --- | --- | --- | --- | --- | --- |
| 1.3 | Registered pregnancies for which the mother received a Mother and Child Protection (MCP) card | -0.13 | 1.44 | -0.09 | 0.93 | 0.93 |
| 2 | Mothers who consumed iron folic acid for 100 days or more when they were pregnant | 0.28 | 1.98 | 0.14 | 0.89 | 0.93 |
| 4.1 | Sex ratio at birth for children born in the last five years | 8.84 | 18.39 | 0.48 | 0.63 | 0.93 |
| 5 | Home births that were conducted by skilled health personnel | -0.52 | 0.58 | -0.89 | 0.38 | 0.81 |
| 6.1 | Children under age 3 years breastfed within one hour of birth | -6.34 | 2.48 | -2.56 | 0.01 | 0.14 |
| 7 | Children under 5 years who are underweight (weight for age) | -3.22 | 3.11 | -1.04 | 0.30 | 0.81 |
| 8.2 | Children with diarrhoea in the 2 weeks preceding the survey who received oral rehydration salts (ORS) | -5.33 | 5.42 | -0.98 | 0.33 | 0.81 |
| 8.3 | Children with diarrhoea in the 2 weeks preceding the survey who received zinc | -1.07 | 3.21 | -0.33 | 0.74 | 0.93 |
| 8.4 | Children with fever or symptoms of ARI in the 2 weeks preceding the survey taken to a health facility or health provide | -6.21 | 5.45 | -1.14 | 0.25 | 0.81 |
| 10.1 | Breastfeeding children age 6-23 months receiving an adequate diet | 0.30 | 1.21 | 0.24 | 0.81 | 0.93 |
| 10.2 | Non-breastfeeding children age 6-23 months receiving an adequate diet | -7.50 | 6.43 | -1.17 | 0.24 | 0.81 |
| 11 | Children age 12-23 months fully vaccinated based on information from either vaccination card or mother's recall | -0.43 | 2.99 | -0.14 | 0.88 | 0.93 |
| 11 | Children age 12-23 months fully vaccinated based on information from vaccination card only | -1.53 | 2.27 | -0.67 | 0.50 | 0.93 |

We would like to mention that our post-intervention NFHS-5 data collection was disrupted due to the pandemic ^4^. Covid-19 is likely to impact the health outcomes, and therefore one may exercise caution while making inferences based on the NFHS-5 data without accounting for the impacts on Covid-19. Unlike previous waves of NFHS data, NFHS-5 data was collected in two phases. The first phase included 22 states before the pandemic (or pre-covid hereafter) and the second phase included the remaining 14 states where the data collection was done during and after the pandemic. Figure 3 maps the pre-covid and post-covid NFHS-5 districts.

**Figure 3.**
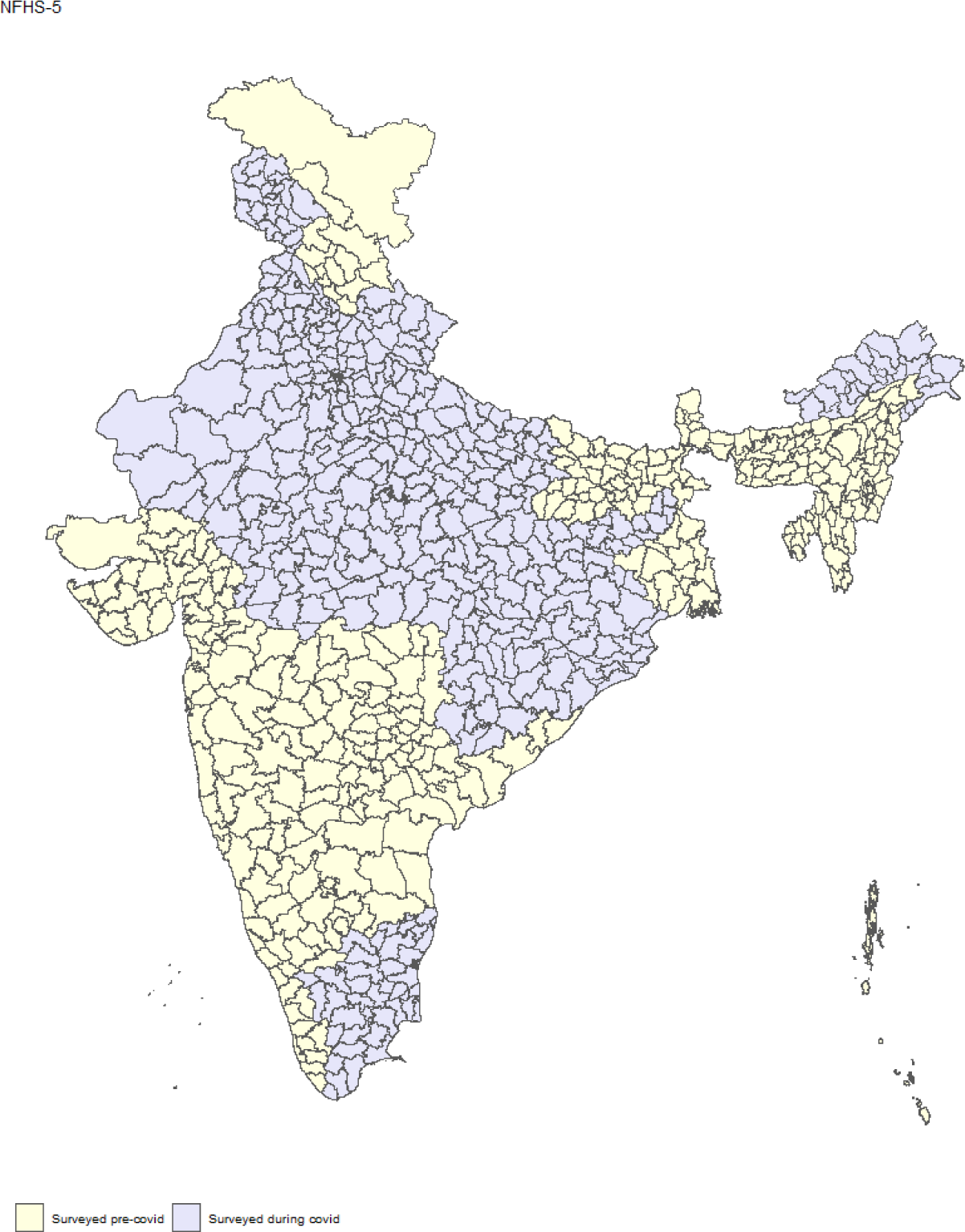
NFHS-5 Survey

Our PSM-DID estimates report the impact of ADP on targeted health outcomes. However, since the estimates are based on full NFHS-5 data for all districts, there could be confounding due to impact of Covid-19 on the targeted health outcomes. Using the full NFHS-5 data including the pre and the post covid data, some of the impacts of the ADP could be masking the impact of Covid-19, confounding the causal estimates of the impact of ADP on targeted health indicators. Moreover, if the impact of the pandemic was felt differently across different districts, this could further bias our causal estimates. Therefore, we implement PSM-DID only on pre-covid data to account for Covid-19 and identify unbiased estimates of ADP on targeted health indicators.

We have a total of 234 pre-covid NFHS-5 districts, of which 40 are ADs. After full matching we are left with 185 control and 28 treatment districts. Table 6 and 7 summarizes the balance of statistics between pre-covid ADs and non-ADs before and after matching respectively. Figure 4 plots the density of observed covariates across ADs and non-ADs before and after matching. The results of the analysis based on pre-covid data are provided in table 8. We find that ADP led to a reduction of around 4% in the percentage of underweight children under 5 years in the ADs, which is statistically significant at 5% level. However, this impact does not survive the FDR when tested using BH procedure as indicated by the adjusted p-value. In addition, we also do not find evidence of any other health outcomes being impacted by the ADP implying that there is no evidence of a reduction in early initiation of breastfeeding that we observed earlier from our PSM-DID estimates using pre and post covid which included all districts.

**Figure 4.**
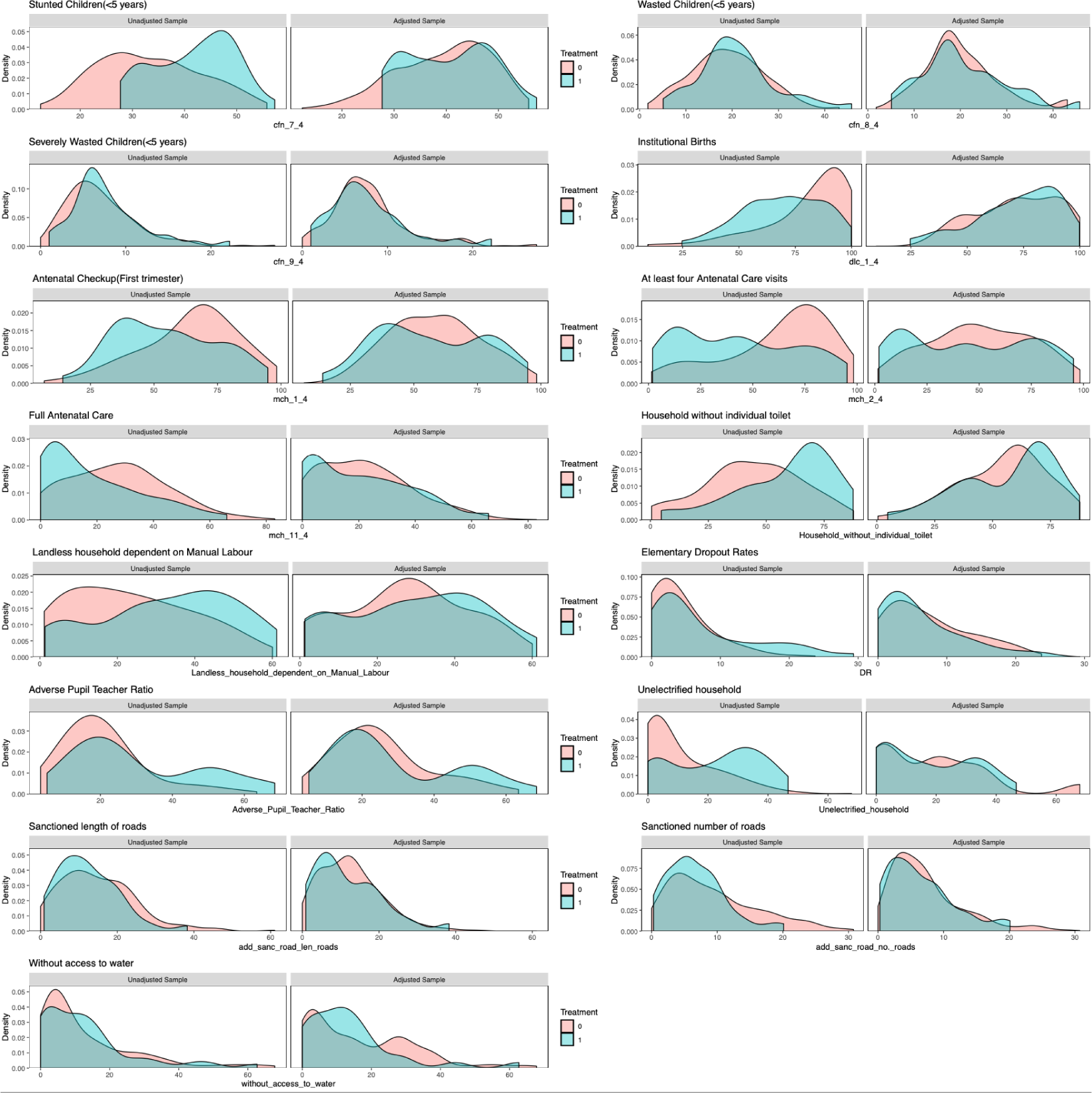
Density plots across AD and non-AD for co-variates before and after matching (Pre-Covid)

**Table 6.**
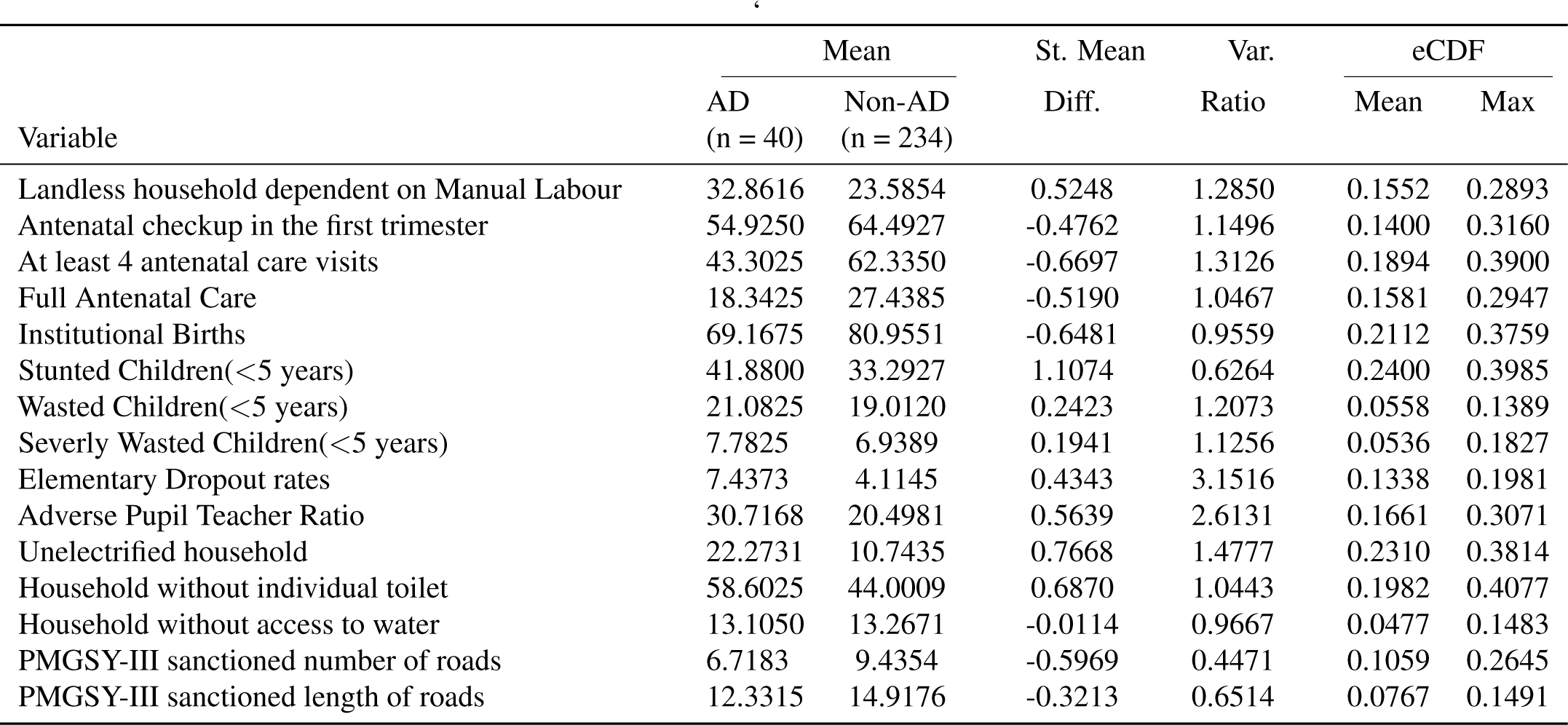
Summary of Covariate Balance before matching (Pre-Covid)

| Variable | Mean |  | St. Mean | Var. | eCDF |  |
| --- | --- | --- | --- | --- | --- | --- |
|  | AD<br>(n = 40) | Non-AD<br>(n = 234) | Diff. | Ratio | Mean | Max |
| Landless household dependent on Manual Labour | 32.8616 | 23.5854 | 0.5248 | 1.2850 | 0.1552 | 0.2893 |
| Antenatal checkup in the first trimester | 54.9250 | 64.4927 | -0.4762 | 1.1496 | 0.1400 | 0.3160 |
| At least 4 antenatal care visits | 43.3025 | 62.3350 | -0.6697 | 1.3126 | 0.1894 | 0.3900 |
| Full Antenatal Care | 18.3425 | 27.4385 | -0.5190 | 1.0467 | 0.1581 | 0.2947 |
| Institutional Births | 69.1675 | 80.9551 | -0.6481 | 0.9559 | 0.2112 | 0.3759 |
| Stunted Children(<5 years) | 41.8800 | 33.2927 | 1.1074 | 0.6264 | 0.2400 | 0.3985 |
| Wasted Children(<5 years) | 21.0825 | 19.0120 | 0.2423 | 1.2073 | 0.0558 | 0.1389 |
| Severly Wasted Children(<5 years) | 7.7825 | 6.9389 | 0.1941 | 1.1256 | 0.0536 | 0.1827 |
| Elementary Dropout rates | 7.4373 | 4.1145 | 0.4343 | 3.1516 | 0.1338 | 0.1981 |
| Adverse Pupil Teacher Ratio | 30.7168 | 20.4981 | 0.5639 | 2.6131 | 0.1661 | 0.3071 |
| Unelectrified household | 22.2731 | 10.7435 | 0.7668 | 1.4777 | 0.2310 | 0.3814 |
| Household without individual toilet | 58.6025 | 44.0009 | 0.6870 | 1.0443 | 0.1982 | 0.4077 |
| Household without access to water | 13.1050 | 13.2671 | -0.0114 | 0.9667 | 0.0477 | 0.1483 |
| PMGSY-III sanctioned number of roads | 6.7183 | 9.4354 | -0.5969 | 0.4471 | 0.1059 | 0.2645 |
| PMGSY-III sanctioned length of roads | 12.3315 | 14.9176 | -0.3213 | 0.6514 | 0.0767 | 0.1491 |

**Table 7.** Summary of Covariate Balance after matching (Pre-Covid)

| Variable | Mean |  | St. Mean | Var. | eCDF |  |
| --- | --- | --- | --- | --- | --- | --- |
|  | AD<br>(n = 28) | Non-AD<br>(n = 185) | Diff. | Ratio | Mean | Max |
| Landless household dependent on Manual Labour | 29.7857 | 27.3705 | 0.1366 | 1.2919 | 0.0464 | 0.2040 |
| Antenatal checkup in the first trimester | 56.3786 | 58.4930 | -0.1052 | 1.5919 | 0.0707 | 0.1953 |
| At least 4 antenatal care visits | 45.8857 | 50.2714 | -0.1543 | 1.5870 | 0.0630 | 0.2384 |
| Full Antenatal Care | 20.6786 | 22.0777 | -0.0798 | 1.2917 | 0.0526 | 0.2009 |
| Institutional Births | 71.9536 | 72.0952 | -0.0078 | 1.0833 | 0.0393 | 0.1117 |
| Stunted Children(<5 years) | 40.7643 | 39.5029 | 0.1627 | 0.9102 | 0.0391 | 0.1302 |
| Wasted Children(<5 years) | 20.6214 | 20.2197 | 0.0470 | 1.3313 | 0.0382 | 0.1064 |
| Severly Wasted Children(<5 years) | 7.7500 | 7.8077 | -0.0133 | 1.1567 | 0.0306 | 0.1399 |
| Elementary Dropout rates | 6.1426 | 7.3270 | -0.1548 | 0.9828 | 0.0689 | 0.1785 |
| Adverse Pupil Teacher Ratio | 29.2537 | 25.4223 | 0.2114 | 1.7883 | 0.0503 | 0.2086 |
| Unelectrified household | 17.6256 | 19.3797 | -0.1167 | 0.6525 | 0.0336 | 0.1003 |
| Household without individual toilet | 56.4786 | 54.1920 | 0.1076 | 1.2597 | 0.0493 | 0.2643 |
| Household without access to water | 13.8464 | 16.6628 | -0.1987 | 0.8222 | 0.0769 | 0.2160 |
| PMGSY-III Sanctioned number of roads | 6.5674 | 7.3315 | -0.1679 | 0.8796 | 0.0458 | 0.1517 |
| PMGSY-III Sanctioned length of roads | 11.9556 | 12.5709 | -0.0765 | 1.2254 | 0.0460 | 0.1531 |

**Table 8.** Results of PSM-DID (Pre-Covid)

| No. | Indicators | Est. | St. err. | t-stat. | p-val. | Adj. p-val. |
| --- | --- | --- | --- | --- | --- | --- |
| 1.3 | Registered pregnancies for which the mother received a Mother and Child Protection (MCP) card | 2.57 | 1.92 | 1.34 | 0.18 | 0.80 |
| 2 | Mothers who consumed iron folic acid for 100 days or more when they were pregnant | 2.94 | 2.93 | 1.00 | 0.32 | 0.80 |
| 4.1 | Sex ratio at birth for children born in the last five years | -23.70 | 37.14 | -0.64 | 0.52 | 0.80 |
| 5 | Home births that were conducted by skilled health personnel | -0.47 | 0.81 | -0.58 | 0.56 | 0.80 |
| 6.1 | Children under age 3 years breastfed within one hour of birth | 1.02 | 3.57 | 0.29 | 0.78 | 0.84 |
| 7 | Children under 5 years who are underweight (weight for age) | -3.87 | 1.95 | -1.99 | 0.05 | 0.62 |
| 8.2 | Children with diarrhoea in the 2 weeks preceding the survey who received oral rehydration salts (ORS) | 7.15 | 9.86 | 0.73 | 0.47 | 0.80 |
| 8.3 | Children with diarrhoea in the 2 weeks preceding the survey who received zinc | 8.69 | 6.60 | 1.32 | 0.19 | 0.80 |
| 8.4 | Children with fever or symptoms of ARI in the 2 weeks preceding the survey taken to a health facility or health provide | -3.26 | 7.19 | -0.45 | 0.65 | 0.80 |
| 10.1 | Breastfeeding children age 6-23 months receiving an adequate diet | -0.79 | 1.90 | -0.41 | 0.68 | 0.80 |
| 10.2 | Non-breastfeeding children age 6-23 months receiving an adequate diet | -2.59 | 5.79 | -0.45 | 0.65 | 0.80 |
| 11 | Children age 12-23 months fully vaccinated based on information from either vaccination card or mother's recall | 0.57 | 3.39 | 0.17 | 0.87 | 0.87 |
| 11 | Children age 12-23 months fully vaccinated based on information from vaccination card only | -2.77 | 4.41 | -0.63 | 0.53 | 0.80 |

### 4.3 Robustness Checks

We run robustness checks for our results using different matching methods on full data that included all districts as well as on the pre-covid districts only. We use two other matching methods that include the Nearest Neighbor Matching (NNM) and the Genetic Matching (GM). NNM pairs every treated unit with the closest eligible control unit, most commonly, the matching is based upon propensity scores. Typically, treated units with the highest propensity scores are paired first. On the other hand,

GM is non-parametric matching, which is a generalization of PS and Mahalanobis distance matching that may include the propensity scores or the observed covariates or both^56,57^. For both methods, we implement matching for 3:1 control versus treatment units.

#### 4.3.1 All districts

We have 97 treated and 291 control districts after matching based on either of the NNM or the GM method using full data that includes all districts. The density plots and covariate balance summary statistics for both methods after matching are included in the appendix (figures S2, S3, and tables S3, S5 respectively). Table 9 reports the PSM-DID results using matching methods NNM and GM. We report results of only those indicators for which we observe a statistically significant impact at conventional levels. The result for all other targeted health outcomes are reported in the appendix (tables S4, S6).

**Table 9.**
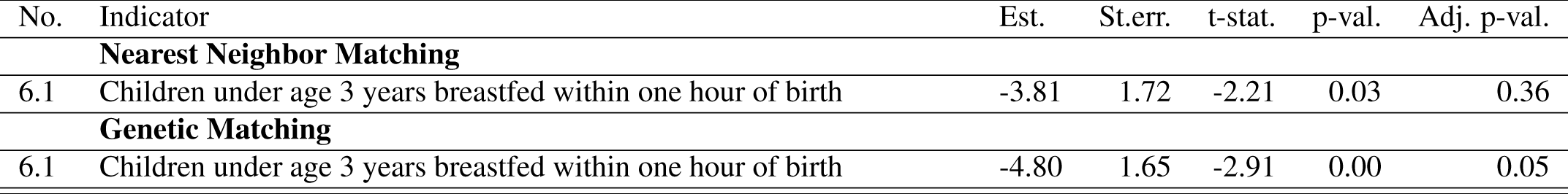
Results from alternative PSM-DID.

PSM-DID estimates from the NNM or the GM methods indicate a reduction of around 4 to 5 percentage points in children under age 3 years breastfed within one hour of birth, which is statistically significant at less than 5% level of significance. The findings from robustness check are in line with the results we get from full matching although the point estimates of PSM-DID based on full matching are higher. In addition, unlike full matching the result under GM also survives FDR following BH procedure, which further indicates that the observed negative outcome is not just by chance.

#### 4.3.2 Pre-covid districts

We have a total of 40 ADs and 234 non-ADs from the pre-covid data that include districts where NFHS-5 data collection was done before covid-19. We run the same set of robustness checks that we implemented for the full data by rerunning our analysis on pre-covid data using NNM and GM. We are left with 30 treated and 90 control districts after matching based on either of the NNM or the GM method. The density plots and covariate balance summary statistics for both methods after matching are included in the appendix (figures S5, S6, and tables S7, S9 respectively). Table 10 reports the PSM-DID results using matching methods NNM and GM on pre-covid data. As previously, we report results of only those indicators for which we observe a statistically significant impact at conventional levels. The result for all other targeted health outcomes are reported in the appendix (tables S8, S10).

**Table 10.**
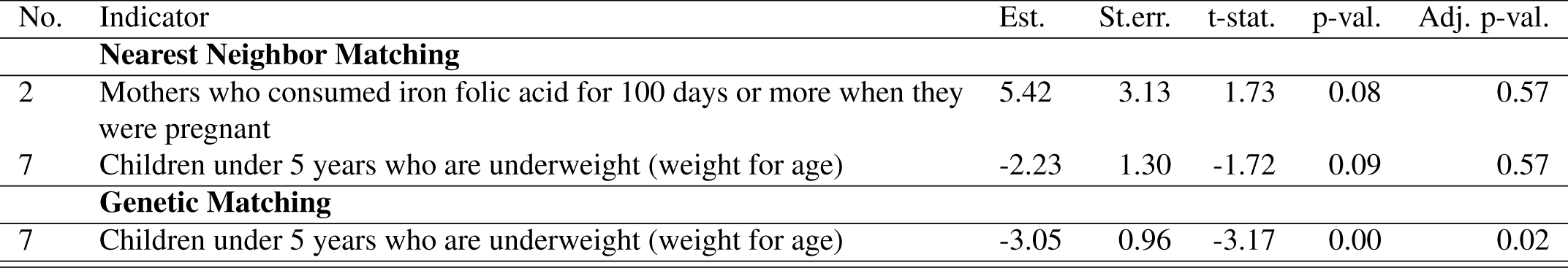
Results from alternative PSM-DID (Pre-covid data only)

DID results post GM indicates that ADP led to a reduction of around 3% points in children under 5 years who are underweight (weight for age) which is statistically significant at less than 1%. Moreover, the result is strong enough to survive the FDR for multiple hypothesis testing using BH test procedure. DID estimates based on NNM too indicate statistically significant reduction in percentage of children under 5 years who are underweight (weight for age) by a little more than 2% though the findings do not survive test for FDR. In addition, under NNM we also find a 5.4% increase in percentage of mothers who had consumed supplemental nutrition in form of iron folic acid for at least 100 days during their pregnancy though it is statistically significant at less than 10% level and does not survive the FDR test. Our results from alternative matching methods are in line with those from full matching as all of them indicate a reduction in the percentage of underweight (weight for age) children younger than 5 years in ADS as an impact of ADP.

## 5 Discussion

Welfare schemes and policies in LMICs are mostly targeted toward addressing severe multidimensional deprivations. Inequality reduction is likely to be an inadvertent but desirable impact of welfare schemes. With this as the stage, ADP in India is a unique of its kind experiment given that it aims to reduce inter-district multidimensional inequalities by identifying the most backward districts in the country, and making additional efforts through ADP so that the ADs catch up with the rest of the country. We evaluate ADP from the lens of health sector. The evaluation metric consists of a composite index, of which health and nutrition indicators carry 30% weightage spanning 31 health data points (over 13 performance indicators) out of a total of 81 data points (over 49 performance indicators). Out of 13 targeted health and nutrition indicators measured using 31 data points, we run an impact evaluation of ADP for 12 indicators that are available in the NFHS and are independent of whether the district would be an AD or not. Prior to our work, assessment of ADP has been limited to documenting the changes or predicting the performance trajectory of ADs compared to non-ADs^34,35,37,58^. Our study is one of the first to estimate the causal impact of ADP on targeted health and nutrition outcomes. We take advantage of the transparent method for identification of an AD and the availability of district-level nationally representative health survey data before and after treatment to estimate the measurable impact of ADP on health and nutrition outcomes.

The NFHS-5 data that we used for our analysis is collected in two phases due to the disruption caused by the Covid-19 pandemic. The first phase is pre-covid that included 22 states and the remaining 14 states are surveyed during the pandemic. Our findings based on full data for all districts indicate that ADP has a negative impact in the ADs by a reduction in early initiation of breastfeeding captured by percentage of children age 3 years who were breastfed within one hour of birth. The summary statistics of the difference in mean between ADs and non-ADs for children under three years breastfed within one hour of birth in table 2 is negative and highly significant for NFHS-5 data though there was no difference in NFHS-4. While the above results could be seen as evidence towards ADP widening the inequality putting ADP in bad light, we could not be confident that these results are casual as we expect that the impact of Covid-19 pandemic are masked in our estimates. COVID-19 pandemic has been characterized by major disruptions in routine health services in both high-income countries and LMICs^59–61^, which has had a profound negative impact on the delivery as well as utilization of Reproductive, Maternal, Newborn, Child Health (RMNCH) services globally as well as in India^62–64^. Multiple studies have found that women who have visited health facilities or have received advice from health workers are more likely to adopt early initiation of breast feeding^65–67^. However, during the pandemic attention of health system including health workers was diverted towards management of Covid-19 that had affected the delivery of health services and availability of health workers for maternal counselling^62,68^. Covid-19 lockdowns and fear of infection further restricted the access to health services. In addition, lack of clear guidelines and recommendations for mothers regarding breastfeeding and new born child care created confusion, which also adversely affected early initiation of breastfeeding^68–70^. Using data from three rounds of NFHS, Mal and Ram (2023)^71^ finds that while early initiation of breast feeding improved significantly over time in India, the estimates from NFHS 2015-16 and NFHS 2019-21 look very similar indicating stagnation in early initiation of breastfeeding. However, it cannot be dismissed that the above findings based on NFHS 2019-21 estimates are uninfluenced by the pandemic given that NFHS data is partly collected during the pandemic. Therefore, we use pre-covid data from NFHS-5 survey to get casual estimates of the impact of ADP which would be unconfounded with the effects of Covid-19. Using the pre-covid data from NFHS-5, we do not find any impact of ADP on early initiation of breast feeding in ADs unlike the negative results uncovered using the full NFHS-5 data.

Our findings from the pre-covid data indicate a meaningful reduction in percentage of children below five years who are underweight. Point estimates from different methods indicate a reduction of 2 to 4% in prevalence of underweight children younger than 5 years. The summary statistics from table 2 indicate that the percentage of underweight children below five years was relatively higher and statistically significant in ADs than non-ADs as measured in NFHS-4. Summary statistics from NFHS-5 full data in table 2 indicate a reduction in percentage of underweight children in both ADs and non-ADs though the difference between ADs and non-ADs persist. Analysis of data based on all five rounds of NFHS starting from 1992-93 to 2019-21 across three decades suggest an overall reduction in the prevalence of underweight children from 56.9% in 1992-93 to 35.5% in 2019-21 with an annual percentage change of −1.6% points per annum^72^. Taking this into account, our estimates of a reduction in 2 to 4% in prevalence of underweight children under 5 years in ADs as an impact of ADP is quite meaningful and encouraging. We rely on estimates from pre-covid data to infer causality indicating that ADP led to a significant reduction in prevalence of underweight children, thus narrowing the gap between ADs and non-ADs. Researchers have expressed their concern regarding child malnutrition in India using pre-covid data as it was observed that 16 out 22 states reported an increase in percentage of underweight children below five years^73,74^. However, our PSM-DID estimates based on pre-covid data reveal that ADP led to a reduction in the percentage of underweight children younger than five years in ADs indicating that ADP could be narrowing the gap between ADs and non-ADs. In addition, there is some weak and non-robust evidence of improvement in uptake of supplemental nutrition among pregnant women indicated by an increase in percentage of mothers who consumed iron folic acid for at least 100 days during pregnancy as can be seen in table 10.

ADP is a multi-sectoral and a multi-component program in India to reduce multi-dimensional inequality between the backward districts and the rest of the country. While ADs are eligible for enhanced funding and priority allocation of various initiatives undertaken by the central and the state governments, the program contributes to ADs by providing a better governance structure and increased accountability through competition among ADs by continuous monitoring and evaluation of their outcomes. ADP provides flexibility to the districts that allow ADs to prioritize sectors to work on, and within the chosen sectors the set of indicators a district would prioritize as targets for improvement. Therefore, it is difficult to pin point specific interventions that would have led to the observed impacts of ADP. However, given the continuous monitoring and evaluation of outcomes, one would expect that districts would grab the low hanging fruits first as also recommended in the operational guidelines of the ADP^39^. Catch-up growth in under-weight children is highly likely if a child gets adequate nutrition since underweight as an indicator of malnutrition is sensitive in the short run, which is achievable in few months^75^. Hence, percentage of underweight children could have been a low hanging fruit in the short run for ADs leading to a reduction in the percentage of underweight children in ADs as an impact of ADP. Using data from earlier rounds of NFHS, researchers have found that lack of dietary diversity, unimproved sanitation, unsafe stool disposal, poor household air quality is positively associated with child underweight in addition to maternal correlates, particularly low BMI^76–78^. India has made significant advancement in Water-Sanitation-Hygiene (WASH) initiatives in the past several years, which could have also helped in contributing to close the gaps. Similarly, the subsidy based Ujjawala scheme for adoption of clean fuel for cooking has led to a rapid transition from polluting fuel to clean cooking fuel that might have helped in improving the household air quality, and subsequently reducing child malnutrition.

We do not find impact of ADP on any other indicator except for reduction in the prevalence of underweight children. There could be several factors that might explain non-impact of ADP on health outcomes. While using the pre-covid NFHS-5^5^ data to assess the impact of ADP, one should not forget that the assessment is based only on one and a half years since the existence of the program. Therefore, it might not be surprising that we don’t see changes in several other indicators. ADs surveyed in the second phase of NFHS-5 survey were exposed to ADP for longer duration, which would make one think that the impacts might be visible in ADs surveyed in the second phase of NFHS-5. However, one runs the risk of confounding with the effects of Covid-19, if one uses data from second phase of NFHS-5 to assess the impact of ADP on ADs. There is no debate that adverse impacts of the pandemic have been felt unequally globally across countries and within countries, which has further exacerbated existing health inequalities^8,79,80^. In the context of ADs, by design most backward districts in the country, predominantly rural regions were identified as ADs. As evident from Table 2, there are significant disparities in different development indicators between ADs and non-ADs. In light of the evidence of the heterogeneous impact of COVID-19 across different socioeconomic groups and regions^63,81–83^, it cannot be ruled out that the rural areas in India were hit harder due to their already stressed health systems even pre-pandemic^84–86^. Therefore, it is possible that findings of a limited positive impact of ADP on health and nutrition outcomes could be masking the gains reversed by the pandemic^87^. It is not unlikely that ADP could have averted several adverse outcomes due to the pandemic. Instead it is comforting to see that there is no evidence of widening of health inequalities between ADs and non-ADs. Lastly, it is also possible that there are visible changes in sectors other than health, which we don’t evaluate in this study. The targeted outcomes of ADP are comprehensive, and are not limited to health and nutrition indicators but include education, agriculture and water resources, financial inclusion and skill development and infrastructure outcomes too. ADP explicitly mentions that the initial focus of ADs should be on achieving low-hanging fruits, which when coupled with competition among ADs could be driving our results of no impact on other health and nutrition indicators^1^. If improvements in targeted health and nutrition outcomes are less visible relative to other targeted sectors over smaller periods of time, competition among ADs might channel efforts and resources toward sectors in which changes are rapidly visible. Therefore, although our findings do not find evidence of significant improvement in health and nutrition indicators, there could be improvement in other non-health indicators as a result of ADP, which is beyond the scope of our research.

We would like to point out few limitations of our study. First, ADP warrants a comprehensive evaluation across all targeted indicators as there might be several interrelated push and pull factors across sectors that need not co-move in the same direction in the short run. Since we evaluate some of the health outcomes from a broader list of targeted outcomes beyond health, we are constrained by our approach to provide multi-sectoral impact evidence of ADP. Secondly, we primarily rely upon pre-pandemic data from phase one of NFHS-5 for our analysis which means that since its implementation, ADs has been exposed to ADP for around one and a half years, which could be a small duration of time for seeing visible impacts of the program. Thirdly, for implementing matching before DID, two out of eleven variables contributing 15% to the composite index for identifying an AD was used from a different data source than the ones used by ADP for identifying AD. This could have a bearing on our counterfactual districts and eventually might impact our DID estimates. We would like to mention that typically program assignment rule is not always known to researchers in quasi-experimental studies, and therefore matching is implemented using variables based on availability of variables at baseline and researchers’ understanding of the problem. In our case, we exploit the transparent mechanism used for identification of ADs under ADP by using the indicators actually used for program assignment for creating counterfactual districts using matching methods, which we believe is a strength of our study. Therefore, even if we use the same variables from a different data source for a couple of variables, we expect them to be strongly correlated with the data used by the ADP. Hence, while there could be small changes, we would not expect major change in findings if we used the same data source as used by ADP for identifying of ADs.

## 6 Conclusion

The Aspirational District Program (ADP) is a unique initiative of the Government of India launched in 2018 that aims to reduce inter-district multi-dimensional inequality. ADP aims to bring the most backward districts to catch up with the rest of the other districts in the country. The program is comprehensive in its scope, as it targets the improvement of several key development indicators spanning health and nutrition, education, agriculture and water resources, financial inclusion and skill development and basic infrastructure. Our results from ADP are encouraging as observed in the short-run with strong evidence of reduction in inter-district inequality through a reduction in the prevalence of underweight children. ADP is an ambitious program that we believe has a modest start towards narrowing down the long persisting gaps between health status of districts in India. It might be too early to make conclusive comment on the success or the failure of ADP. Using NFHS 4 and 5 data, Subramanian et al. (2023)^36^ assessed the progress of Indian districts in terms of achievement of 33 Sustainable Development Goal (SDG) indicators by 2030. Their findings suggest no clear pattern that AD’s are more likely to meet the SDG targets compared to other districts though they admit the possibility of Covid-19 related disruptions during the intervening survey periods could have influenced the outcomes. While Subramanian et al. (2023)^36^ strongly laud ADP as a right step towards achieving of SDGs and reducing inter-district inequality, they recommend that multiple separate ADPs for different indicators might be more effective as it would improve targeting instead of the existing comprehensive multi-sectoral ADP. We appreciate their recommendations, yet we feel that ADP must be given more time primarily because of the Covid-19 disruptions that have impacted their functioning as well as evaluation. Round 6 of NFHS data collection between 2023-2024 is underway, which might be able to provide us with a better picture on the performance of ADP. Using micro-level data recent research has drawn attention to heterogeneity in health and nutrition indicators at sub-district level even at the level of villages based on which researchers have argued for geographical targeting at micro-spatial units^20–22^. This provides an opportunity for a program like ADP to capitalize on these research findings by identifying the micro hotspots and targeting these small spatial units within a district for potential improvement. Under ADP, ADs have flexibility in terms of choosing the priority sector and provision to tailor interventions as suitable to local contexts, which could be leveraged based on findings from sub-district analysis of health outcomes. We suggest that future research efforts should be made toward impact evaluation of all the targeted indicators using post-covid data, which will provide a comprehensive unbiased evaluation of ADP.

## Data Availability

All data produced in the present study are available upon reasonable request to the authors

## Acknowledgements

We are thankful to J V Meenakshi, Ram Singh, Sangeeta Bansal and participants at the Annual Economics Conference 2022 organized at IISER Bhopal for their useful comments and feedback on an earlier version of the paper.

## Author contributions statement

SKA conceived the project, SM prepared the data and conducted the analysis under supervision of SKA, SM wrote the first draft of the paper, SKA prepared the current draft. All authors reviewed the manuscript. (Sandip K. Agarwal (SKA); Shubham Mishra (SM))

## Additional information

### Data availability

The datasets used and/or analysed during the current study available from the corresponding author on reasonable request.

## Supplementary Appendix

### 1 ADP: Health Indicators

Table S1 below lists the 31 health indicators across 11 core health variables targeted for improvement in ADP (measured in percentages unless specified) along with the corresponding NFHS variable.

**Table S1:**
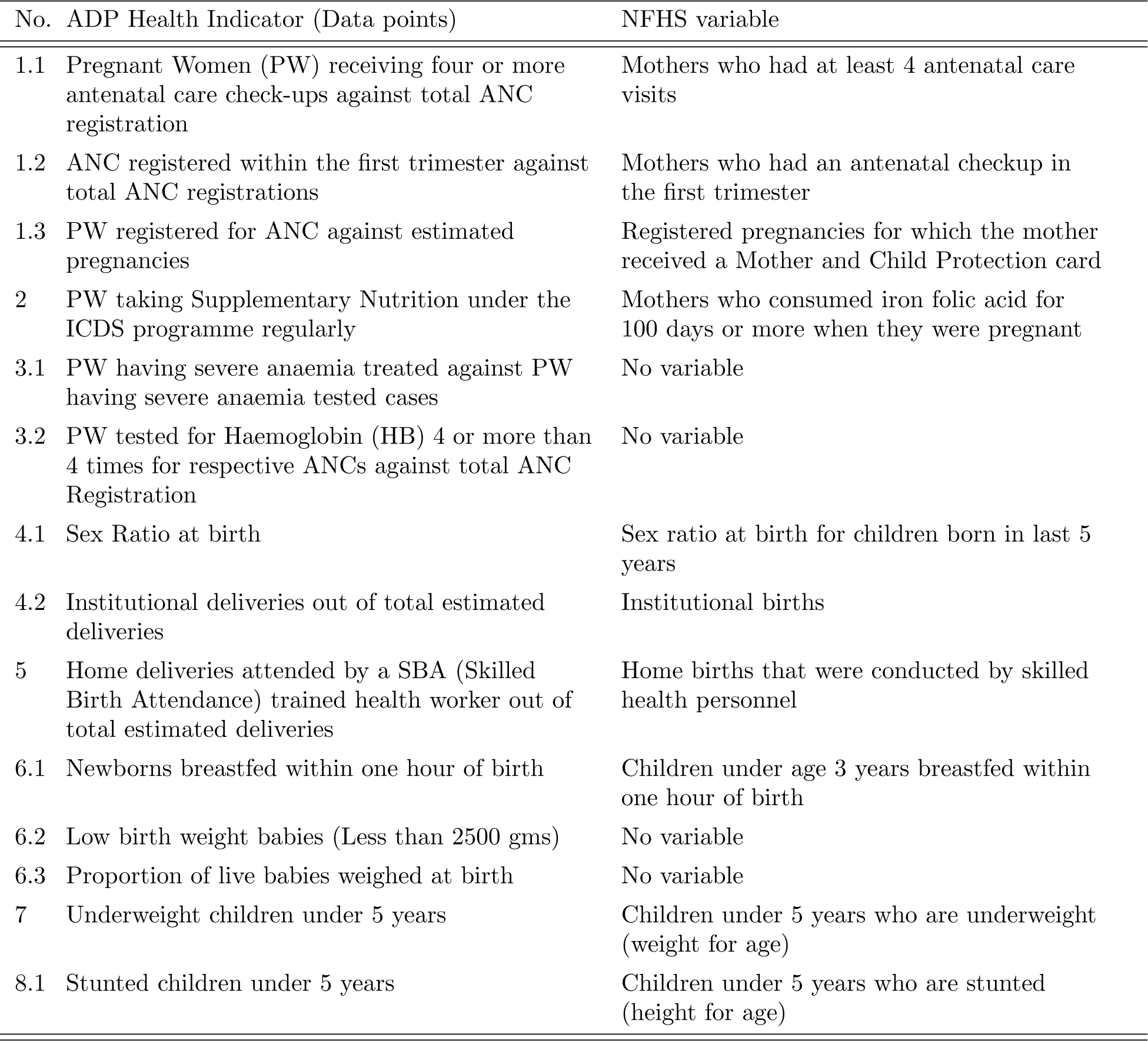
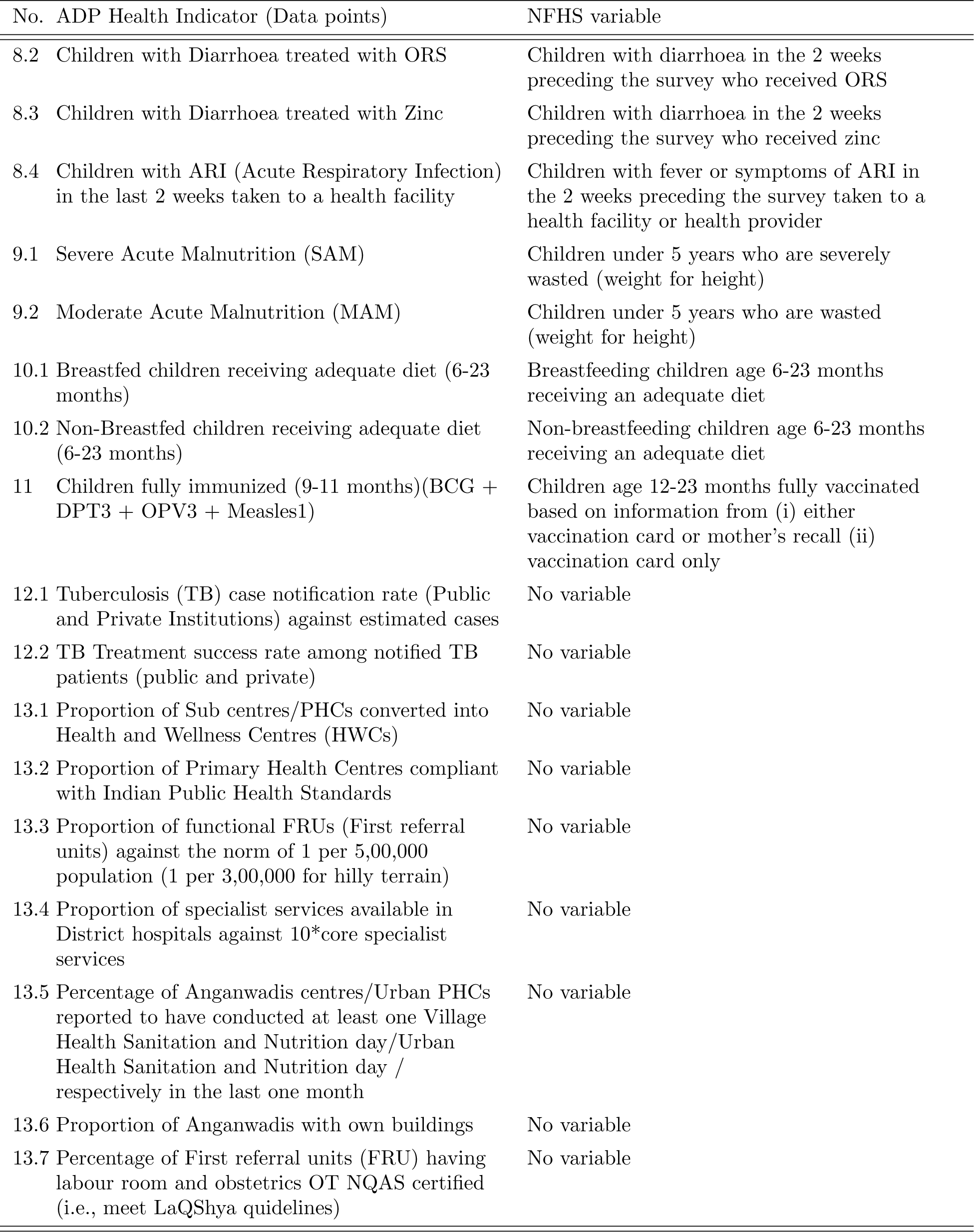
List of Health indicators (data points) targeted by ADP (Source: Operational Guidelines for improving Health and Nutrition Status in Aspirational Districts - Annexure 1.

| No. | ADP Health Indicator (Data points) | NFHS variable |
| --- | --- | --- |
| 1.1 | Pregnant Women (PW) receiving four or more antenatal care check-ups against total ANC registration | Mothers who had at least 4 antenatal care visits |
| 1.2 | ANC registered within the first trimester against total ANC registrations | Mothers who had an antenatal checkup in the first trimester |
| 1.3 | PW registered for ANC against estimated pregnancies | Registered pregnancies for which the mother received a Mother and Child Protection card |
| 2 | PW taking Supplementary Nutrition under the ICDS programme regularly | Mothers who consumed iron folic acid for 100 days or more when they were pregnant |
| 3.1 | PW having severe anaemia treated against PW having severe anaemia tested cases | No variable |
| 3.2 | PW tested for Haemoglobin (HB) 4 or more than 4 times for respective ANCs against total ANC Registration | No variable |
| 4.1 | Sex Ratio at birth | Sex ratio at birth for children born in last 5 years |
| 4.2 | Institutional deliveries out of total estimated deliveries | Institutional births |
| 5 | Home deliveries attended by a SBA (Skilled Birth Attendance) trained health worker out of total estimated deliveries | Home births that were conducted by skilled health personnel |
| 6.1 | Newborns breastfed within one hour of birth | Children under age 3 years breastfed within one hour of birth |
| 6.2 | Low birth weight babies (Less than 2500 gms) | No variable |
| 6.3 | Proportion of live babies weighed at birth | No variable |
| 7 | Underweight children under 5 years | Children under 5 years who are underweight (weight for age) |
| 8.1 | Stunted children under 5 years | Children under 5 years who are stunted (height for age) |

Table S1: List of Health indicators (data points) targeted by ADP (Source: Operational Guidelines for improving Health and Nutrition Status in Aspirational Districts - Annexure 1 (Continued))
| No. | ADP Health Indicator (Data points) | NFHS variable |
| --- | --- | --- |
| 8.2 | Children with Diarrhoea treated with ORS | Children with diarrhoea in the 2 weeks preceding the survey who received ORS |
| 8.3 | Children with Diarrhoea treated with Zinc | Children with diarrhoea in the 2 weeks preceding the survey who received zinc |
| 8.4 | Children with ARI (Acute Respiratory Infection) in the last 2 weeks taken to a health facility | Children with fever or symptoms of ARI in the 2 weeks preceding the survey taken to a health facility or health provider |
| 9.1 | Severe Acute Malnutrition (SAM) | Children under 5 years who are severely wasted (weight for height) |
| 9.2 | Moderate Acute Malnutrition (MAM) | Children under 5 years who are wasted (weight for height) |
| 10.1 | Breastfed children receiving adequate diet (6-23 months) | Breastfeeding children age 6-23 months receiving an adequate diet |
| 10.2 | Non-Breastfed children receiving adequate diet (6-23 months) | Non-breastfeeding children age 6-23 months receiving an adequate diet |
| 11 | Children fully immunized (9-11 months)(BCG + DPT3 + OPV3 + Measles1) | Children age 12-23 months fully vaccinated based on information from (i) either vaccination card or mother's recall (ii) vaccination card only |
| 12.1 | Tuberculosis (TB) case notification rate (Public and Private Institutions) against estimated cases | No variable |
| 12.2 | TB Treatment success rate among notified TB patients (public and private) | No variable |
| 13.1 | Proportion of Sub centres/PHCs converted into Health and Wellness Centres (HWCs) | No variable |
| 13.2 | Proportion of Primary Health Centres compliant with Indian Public Health Standards | No variable |
| 13.3 | Proportion of functional FRUs (First referral units) against the norm of 1 per 5,00,000 population (1 per 3,00,000 for hilly terrain) | No variable |
| 13.4 | Proportion of specialist services available in District hospitals against 10*core specialist services | No variable |
| 13.5 | Percentage of Anganwadis centres/Urban PHCs reported to have conducted at least one Village Health Sanitation and Nutrition day/Urban Health Sanitation and Nutrition day / respectively in the last one month | No variable |
| 13.6 | Proportion of Anganwadis with own buildings | No variable |
| 13.7 | Percentage of First referral units (FRU) having labour room and obstetrics OT NQAS certified (i.e., meet LaQShya guidelines) | No variable |

Table S2 below lists the health indicators from NFHS data that we included in our evaluation along with their definitions.

**Table S2:** Definition of NFHS variables included in the analysis for evaluation.

| No. NFHS variable | Definition |
| --- | --- |
| 1.3 Registered pregnancies for which the mother received a Mother and Child Protection (MCP) card | Percentage of pregnancies that were registered for which the mother received a Mother and Child Protection (MCP) card |
| 2 Mothers who consumed iron folic acid for 100 days or more when they were pregnant | Percentage of Mothers who consumed iron folic acid for 100 days or more when they were pregnant |
| 4.1 Sex ratio at birth for children born in last 5 years | Sex ratio at birth for children born in the last five years (females per 1,000 males) |
| 5 Home births that were conducted by skilled health personnel | Percentage of home births that were conducted by Doctor / nurse / LHV / ANM / midwife / other health personnel. |
| 6.1 Children under age 3 years breastfed within one hour of birth | Percentage of children under age 3 years breastfed within one hour of birth based on the last child born in the 3 years before the survey |
| 7 Children under 5 years who are underweight (weight for age) | Percentage of children under 5 years who are underweight (weight-for-age), where below -2 standard deviations is underweight based on the WHO standard. |
| 8.2 Children with diarrhoea in the 2 weeks preceding the survey who received ORS | Percentage of children with diarrhoea in the 2 weeks preceding the survey who received oral rehydration salts (ORS) |
| 8.3 Children with diarrhoea in the 2 weeks preceding the survey who received zinc | Percentage of children with diarrhoea in the 2 weeks preceding the survey who received zinc |
| 8.4 Children with fever or symptoms of ARI in the 2 weeks preceding the survey taken to a health facility or health provider | Percentage of children with fever or symptoms of ARI in the 2 weeks preceding the survey taken to a health facility or health provider |
| 10.1 Breastfeeding children age 6-23 months receiving an adequate diet | Percentage of breastfeeding children age 6-23 months receiving an adequate diet based on the youngest child living with the mother. Adequate diet is breastfed children receiving 4 or more food groups and a minimum meal frequency, that is, receiving solid or semi-solid food at least twice a day for breastfed infants 6-8 months and at least three times a day for breastfed children 9-23 months |

Table S2: Definition of NFHS variables included in the analysis for evaluation (Continued)
| No. NFHS variable | Definition |
| --- | --- |
| 10.2 Non-breastfeeding children age 6-23 months receiving an adequate diet | Percentage of non-breastfeeding children age 6-23 months receiving an adequate diet based on the youngest child living with the mother. Adequate diet is non-breastfed children fed with a minimum of 3 infant and young Child feeding practices (fed with other milk or milk products at least twice a day, a minimum meal frequency that is, solid or semi-solid foods from at least four food groups not including the milk or milk products food group) |
| 11 Children age 12-23 months fully vaccinated based on information from (i) either vaccination card or mother's recall (ii) vaccination card only | Percentage of children vaccinated with BCG, measles-containing vaccine (MCV) / MR/ MMR/ Measles, and 3 doses each of polio (excluding polio vaccine given at birth) and DPT or penta vaccine. |

### 2 PSM (Full data / All districts)

#### 2.1 Full matching

**Figure S1:**
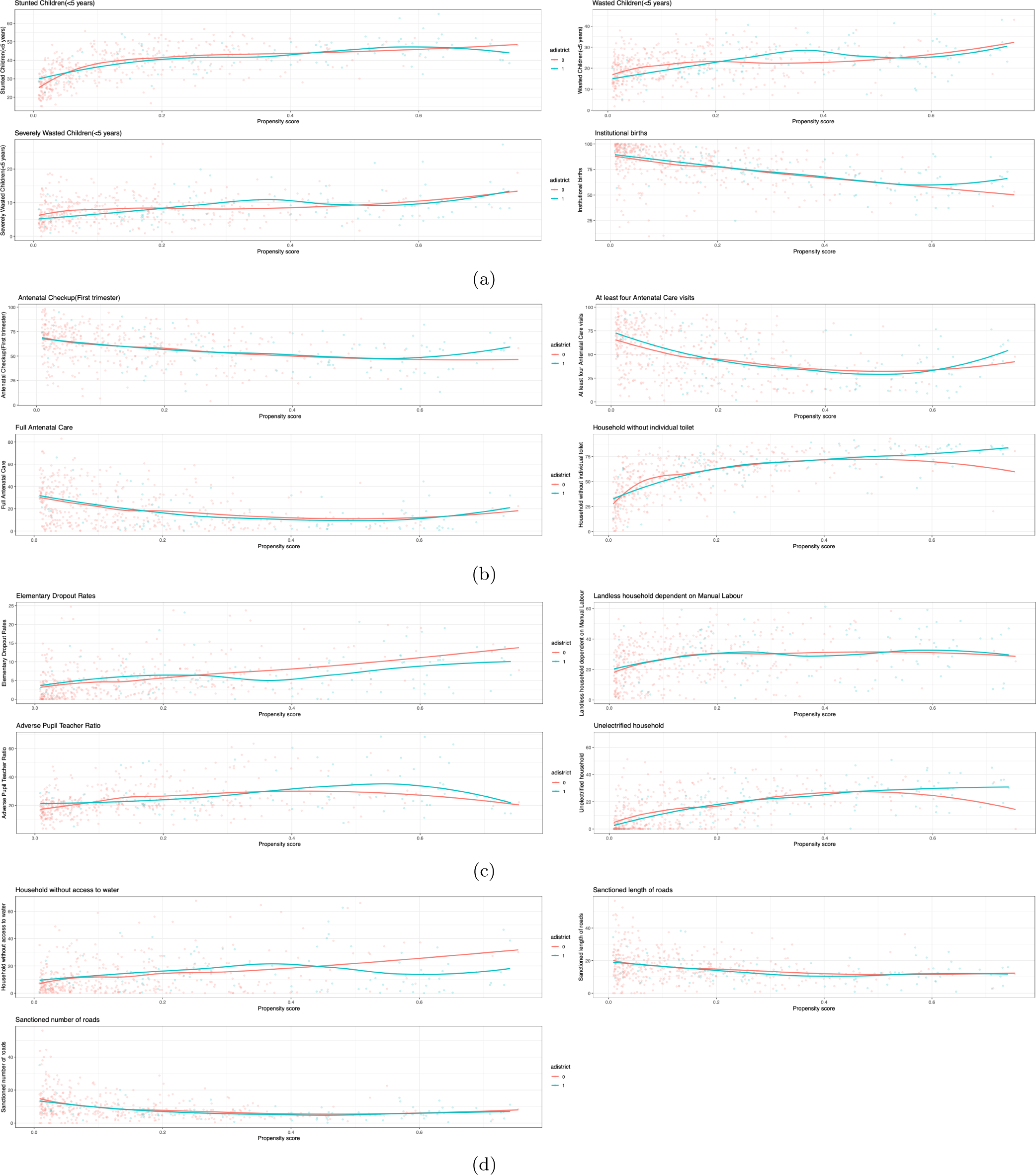
Scatter plot of estimated propensity scores across different covariates

#### 2.2 Nearest Neigbour Matching

**Figure S2:**
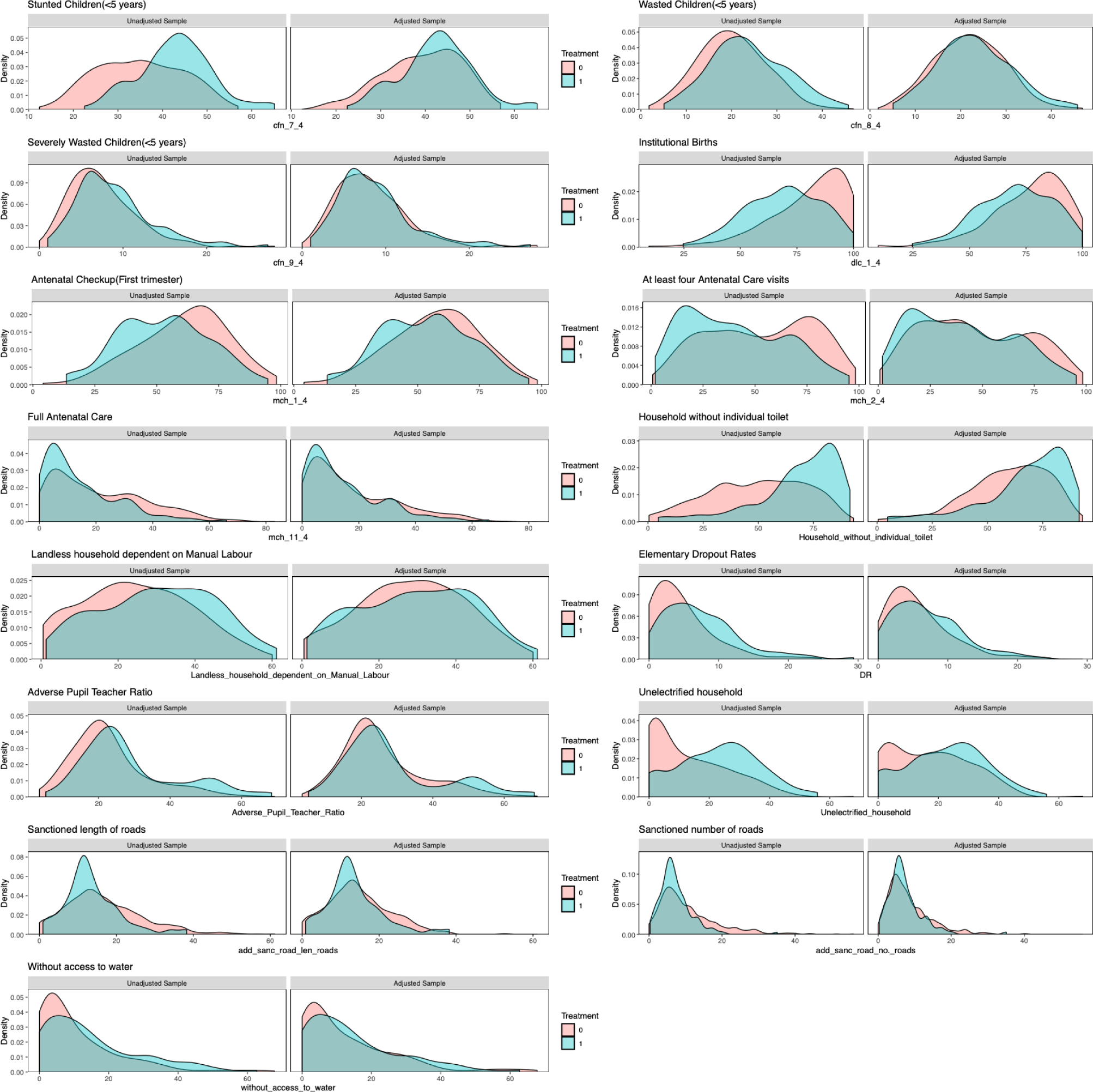
Balance of covariate density plots for Propensity Score matched samples using Nearest Neighbor Matching

**Table S3:** Summary of Covariate Balance after matching (Nearest Neighbour)

| Variable | Mean |  | St. Mean | Var. | eCDF |  |
| --- | --- | --- | --- | --- | --- | --- |
|  | AD<br>(n = 97) | Non-AD<br>(n = 291) | Diff. | Ratio | Mean | Max |
| Landless household dependent on Manual Labour | 29.7683 | 28.4953 | 0.0856 | 1.1852 | 0.0346 | 0.0928 |
| Antenatal checkup in the first trimester | 53.3031 | 57.2395 | -0.2186 | 1.0396 | 0.0591 | 0.1375 |
| At least 4 antenatal care visits | 40.4536 | 46.5577 | -0.2547 | 0.9160 | 0.0695 | 0.1271 |
| Full Antenatal Care | 14.8268 | 18.0186 | -0.2365 | 0.7714 | 0.0581 | 0.1065 |
| Institutional Births | 69.9948 | 76.4409 | -0.3856 | 1.0052 | 0.1131 | 0.2371 |
| Stunted Children(<5 years) | 42.6680 | 39.3790 | 0.3999 | 0.8944 | 0.0812 | 0.1821 |
| Wasted Children(<5 years) | 23.2505 | 21.9584 | 0.1534 | 1.1655 | 0.0356 | 0.0859 |
| Severly Wasted Children(<5 years) | 8.6979 | 8.1368 | 0.1201 | 1.2079 | 0.0277 | 0.0756 |
| Elementary Dropout rates | 7.0536 | 5.7576 | 0.2335 | 1.1714 | 0.0800 | 0.1718 |
| Adverse Pupil Teacher Ratio | 28.8265 | 25.9692 | 0.2040 | 1.5792 | 0.0492 | 0.1203 |
| Unelectrified household | 22.5025 | 17.3067 | 0.3863 | 1.0100 | 0.1079 | 0.2062 |
| Household without individual toilet | 67.2814 | 59.2722 | 0.4179 | 1.1437 | 0.1119 | 0.2543 |
| Household without access to water | 14.8103 | 12.8625 | 0.1397 | 0.9758 | 0.0562 | 0.1478 |
| PMGSY-III Sanctioned number of roads | 7.2737 | 8.3721 | -0.2280 | 0.6668 | 0.0502 | 0.1237 |
| PMGSY-III Sanctioned length of roads | 13.6467 | 14.8539 | -0.1643 | 0.7990 | 0.0565 | 0.1340 |

**Table S4:**
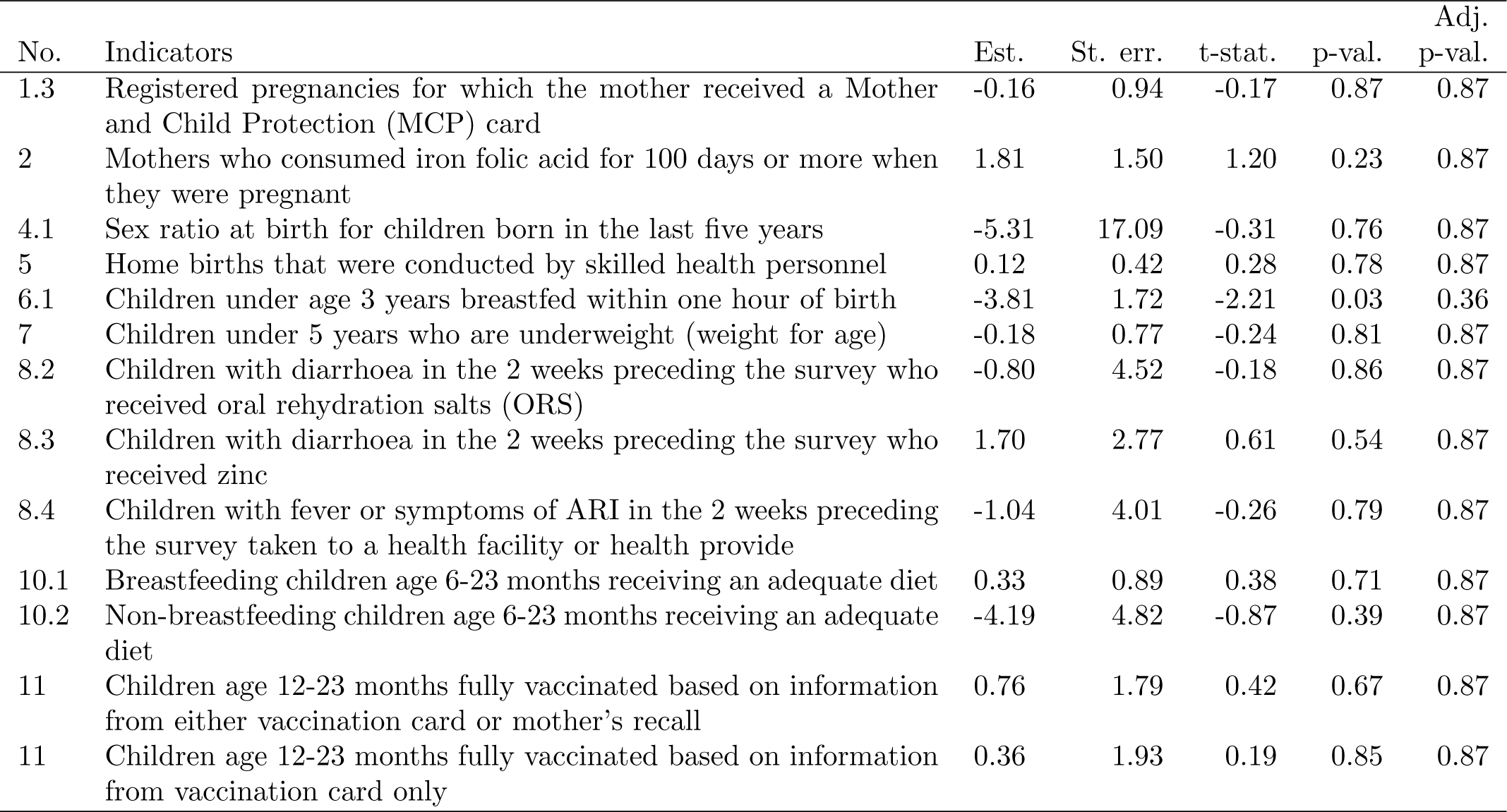
Results of PSM-DID (Nearest Neighbour)

#### 2.3 Genetic Matching

**Figure S3:**
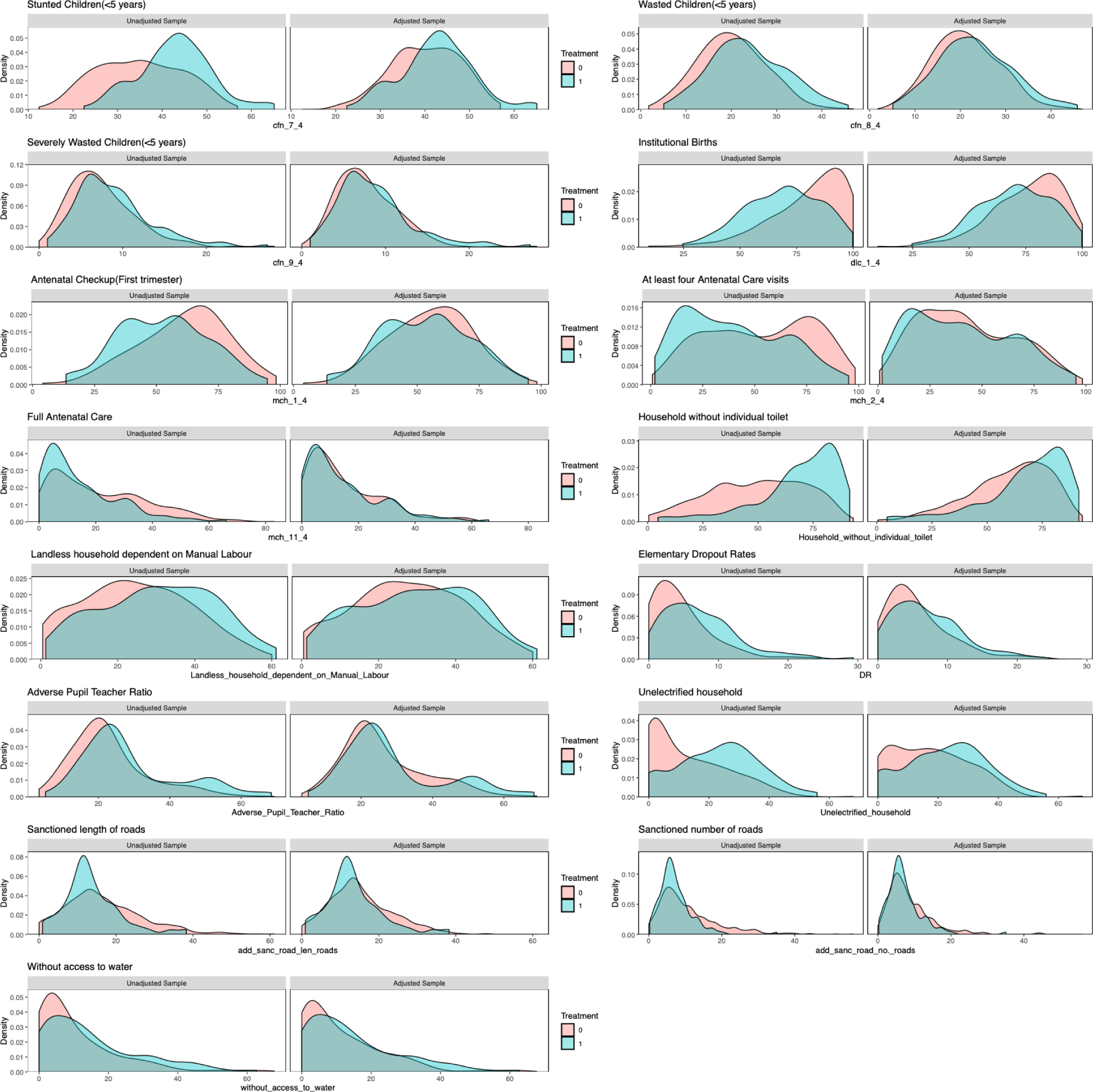
Balance of covariate density plots for matched samples using Genetic Matching

**Table S5:**
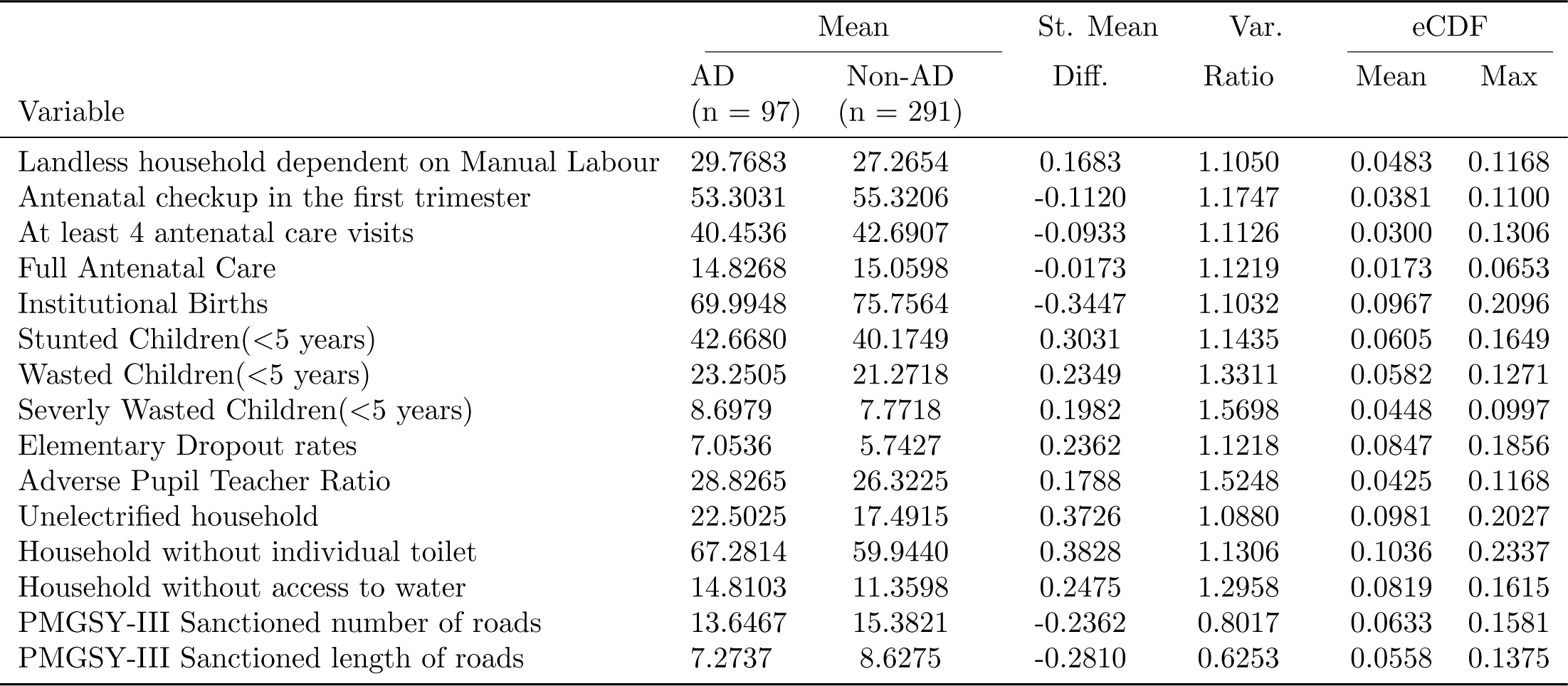
Summary of Covariate Balance after matching (Genetic Matching)

**Table S6:** Results of PSM-DID (Genetic Matching)

| No. | Indicators | Est. | St. err. | t-stat. | p-val. | Adj.<br>p-val. |
| --- | --- | --- | --- | --- | --- | --- |
| 1.3 | Registered pregnancies for which the mother received a Mother and Child Protection (MCP) card | -0.54 | 0.81 | -0.67 | 0.51 | 0.95 |
| 2 | Mothers who consumed iron folic acid for 100 days or more when they were pregnant | 2.05 | 1.29 | 1.60 | 0.11 | 0.72 |
| 4.1 | Sex ratio at birth for children born in the last five years | -4.18 | 17.99 | -0.23 | 0.82 | 0.95 |
| 5 | Home births that were conducted by skilled health personnel | 0.10 | 0.38 | 0.26 | 0.80 | 0.95 |
| 6.1 | Children under age 3 years breastfed within one hour of birth | -4.80 | 1.65 | -2.91 | 0.00 | 0.05 |
| 7 | Children under 5 years who are underweight (weight for age) | -0.16 | 0.67 | -0.24 | 0.81 | 0.95 |
| 8.2 | Children with diarrhoea in the 2 weeks preceding the survey who received oral rehydration salts (ORS) | -3.43 | 3.97 | -0.86 | 0.39 | 0.95 |
| 8.3 | Children with diarrhoea in the 2 weeks preceding the survey who received zinc | 0.18 | 2.35 | 0.08 | 0.94 | 0.95 |
| 8.4 | Children with fever or symptoms of ARI in the 2 weeks preceding the survey taken to a health facility or health provide | 0.45 | 3.38 | 0.13 | 0.90 | 0.95 |
| 10.1 | Breastfeeding children age 6-23 months receiving an adequate diet | 0.05 | 0.83 | 0.06 | 0.95 | 0.95 |
| 10.2 | Non-breastfeeding children age 6-23 months receiving an adequate diet | -4.58 | 4.06 | -1.13 | 0.26 | 0.95 |
| 11 | Children age 12-23 months fully vaccinated based on information from either vaccination card or mother's recall | -0.16 | 1.77 | -0.09 | 0.93 | 0.95 |
| 11 | Children age 12-23 months fully vaccinated based on information from vaccination card only | -1.11 | 1.90 | -0.58 | 0.56 | 0.95 |

### 3 PSM (Pre-Covid Data)

#### 3.1 Full matching method

**Figure S4:**
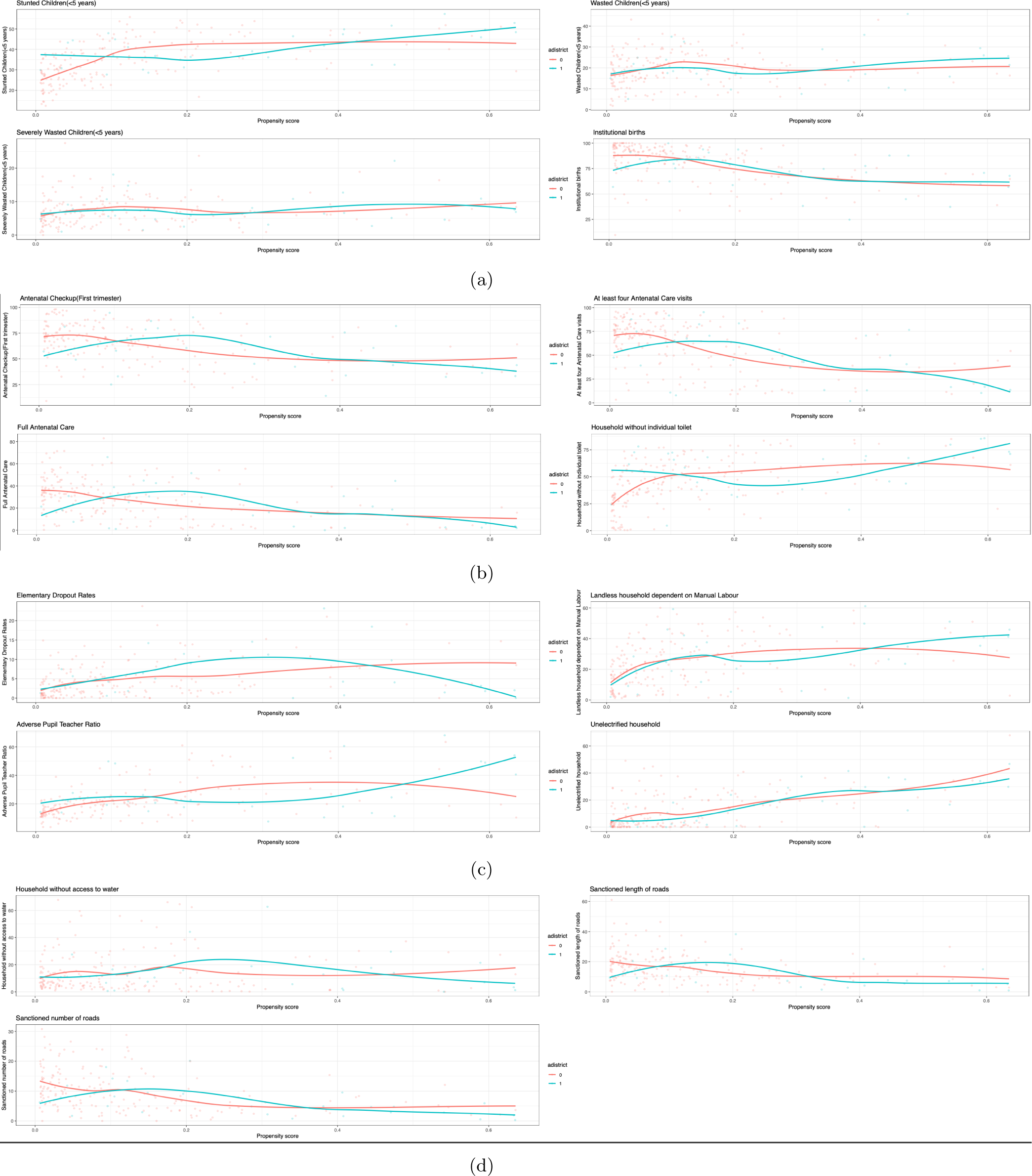
Scatter plot of estimated propensity scores across different covariates using Pre-covid data

#### 3.2 Nearest Neighbour Matching

**Figure S5:**
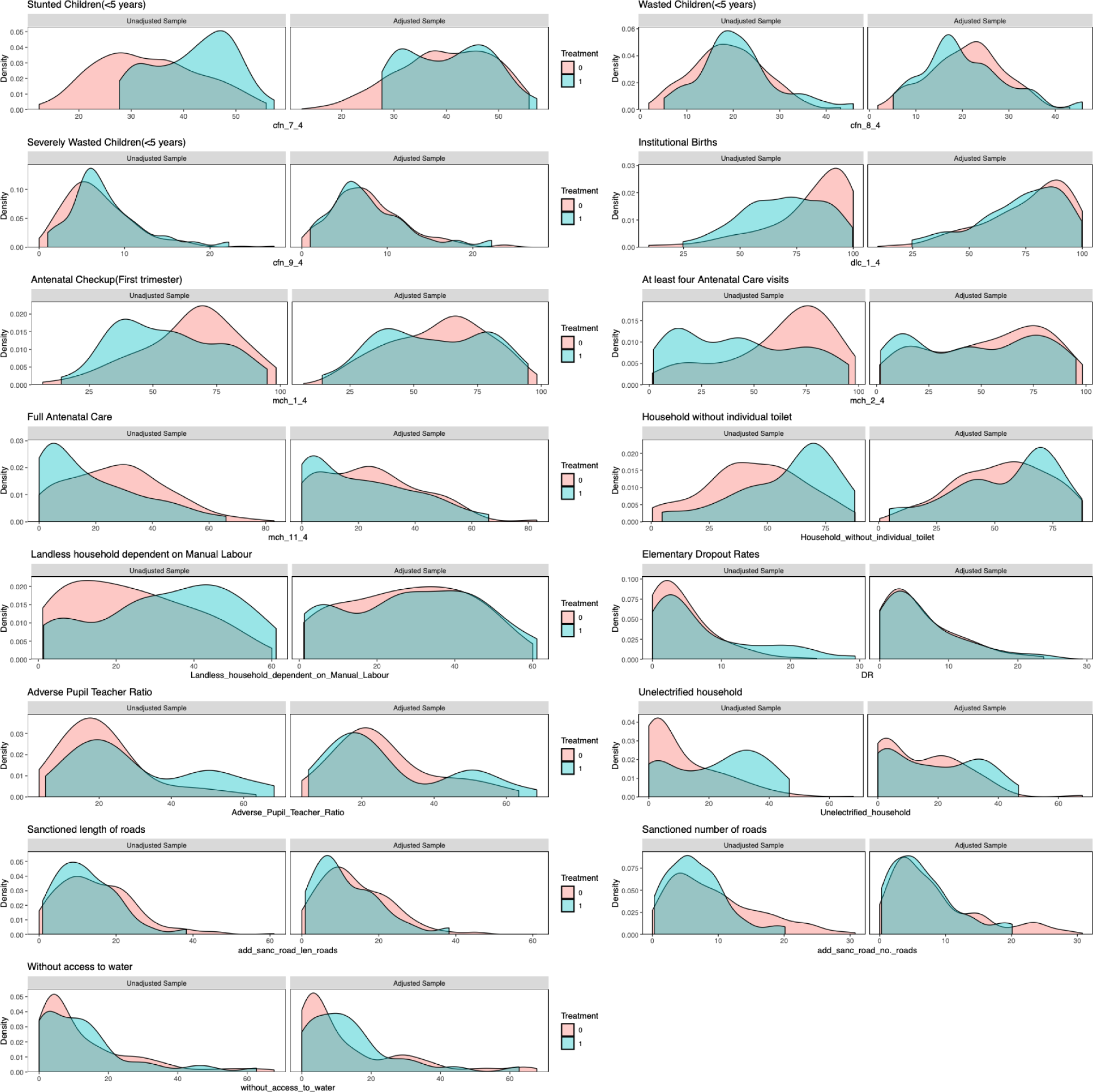
Balance of covariate density plots for Propensity Score matched samples using Nearest Neighbor Matching

**Table S7:** Summary of Covariate Balance after matching (Nearest Neighbour)

| Variable | Mean |  | St. Mean | Var. | eCDF |  |
| --- | --- | --- | --- | --- | --- | --- |
|  | AD<br>(n = 30) | Non-AD<br>(n = 90) | Diff. | Ratio | Mean | Max |
| Landless household dependent on Manual Labour | 28.9157 | 28.0767 | 0.0475 | 1.2511 | 0.0330 | 0.0778 |
| Antenatal checkup in the first trimester | 57.5400 | 59.8633 | -0.1156 | 1.3199 | 0.0551 | 0.1444 |
| At least 4 antenatal care visits | 47.8700 | 53.2544 | -0.1895 | 1.2494 | 0.0538 | 0.1667 |
| Full Antenatal Care | 21.3767 | 24.4044 | -0.1727 | 1.1408 | 0.0574 | 0.1667 |
| Institutional Births | 72.8500 | 76.0967 | -0.1785 | 1.0714 | 0.0603 | 0.1444 |
| Stunted Children(<5 years) | 40.5400 | 39.8933 | 0.0834 | 0.8568 | 0.0320 | 0.0889 |
| Wasted Children(<5 years) | 20.5067 | 20.9689 | -0.0541 | 1.3012 | 0.0469 | 0.1778 |
| Severly Wasted Children(<5 years) | 7.8067 | 7.7711 | 0.0082 | 1.2133 | 0.0228 | 0.0778 |
| Elementary Dropout rates | 5.9952 | 5.7442 | 0.0328 | 1.1973 | 0.0262 | 0.0889 |
| Adverse Pupil Teacher Ratio | 27.9035 | 26.1635 | 0.0960 | 1.6603 | 0.0542 | 0.1667 |
| Unelectrified household | 18.4323 | 14.5868 | 0.2558 | 1.2403 | 0.0726 | 0.2333 |
| Household without individual toilet | 54.6300 | 53.0433 | 0.0746 | 1.2804 | 0.0495 | 0.2000 |
| Household without access to water | 14.2567 | 13.5722 | 0.0483 | 0.7445 | 0.0821 | 0.2444 |
| PMGSY-III Sanctioned number of roads | 6.5173 | 8.1677 | -0.3625 | 0.5577 | 0.0666 | 0.1667 |
| PMGSY-III Sanctioned length of roads | 11.7417 | 13.7467 | -0.2491 | 0.9604 | 0.0740 | 0.1889 |

**Table S8:** Results of PSM-DID (Nearest Neighbour Matching)

| No. | Indicators | Est. | St. err. | t-stat. | p-val. | Adj.<br>p-val. |
| --- | --- | --- | --- | --- | --- | --- |
| 1.3 | Registered pregnancies for which the mother received a Mother and Child Protection (MCP) card | 0.70 | 2.21 | 0.32 | 0.75 | 0.97 |
| 2 | Mothers who consumed iron folic acid for 100 days or more when they were pregnant | 5.42 | 3.13 | 1.73 | 0.08 | 0.57 |
| 4.1 | Sex ratio at birth for children born in the last five years | -1.23 | 34.77 | -0.04 | 0.97 | 0.97 |
| 5 | Home births that were conducted by skilled health personnel | 0.05 | 0.79 | 0.06 | 0.95 | 0.97 |
| 6.1 | Children under age 3 years breastfed within one hour of birth | 3.37 | 3.53 | 0.96 | 0.34 | 0.93 |
| 7 | Children under 5 years who are underweight (weight for age) | -2.23 | 1.30 | -1.72 | 0.09 | 0.57 |
| 8.2 | Children with diarrhoea in the 2 weeks preceding the survey who received oral rehydration salts (ORS) | -0.68 | 8.81 | -0.08 | 0.94 | 0.97 |
| 8.3 | Children with diarrhoea in the 2 weeks preceding the survey who received zinc | 2.43 | 5.27 | 0.46 | 0.65 | 0.93 |
| 8.4 | Children with fever or symptoms of ARI in the 2 weeks preceding the survey taken to a health facility or health provide | -4.61 | 7.18 | -0.64 | 0.52 | 0.93 |
| 10.1 | Breastfeeding children age 6-23 months receiving an adequate diet | -2.04 | 1.57 | -1.30 | 0.20 | 0.87 |
| 10.2 | Non-breastfeeding children age 6-23 months receiving an adequate diet | -3.09 | 4.45 | -0.69 | 0.49 | 0.93 |
| 11 | Children age 12-23 months fully vaccinated based on information from either vaccination card or mother's recall | 1.86 | 2.78 | 0.67 | 0.50 | 0.93 |
| 11 | Children age 12-23 months fully vaccinated based on information from vaccination card only | -1.69 | 3.54 | -0.48 | 0.63 | 0.93 |

#### 3.3 Genetic Matching

**Figure S6:**
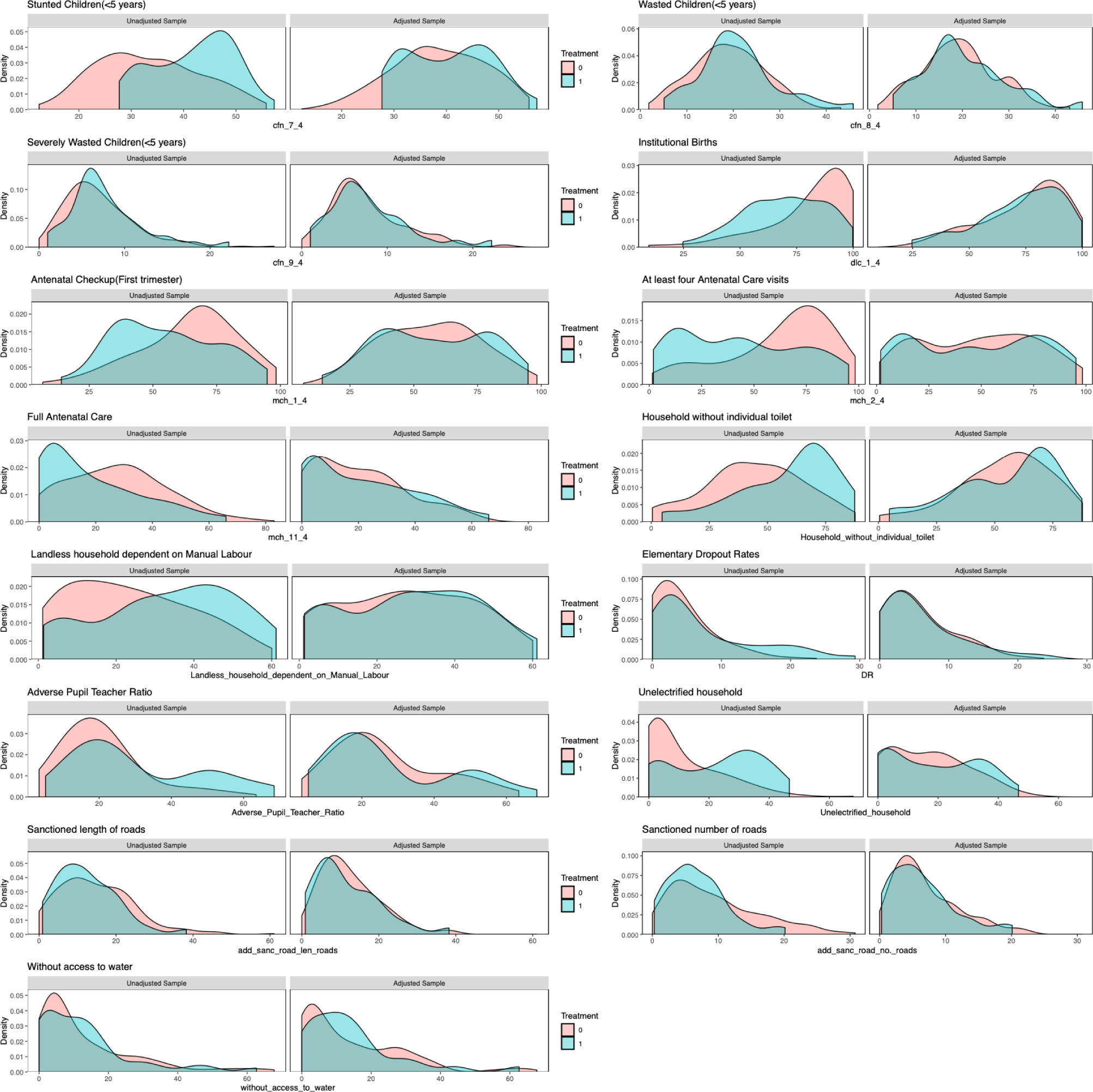
Balance of covariate density plots for matched samples using Genetic Matching

**Table S9:**
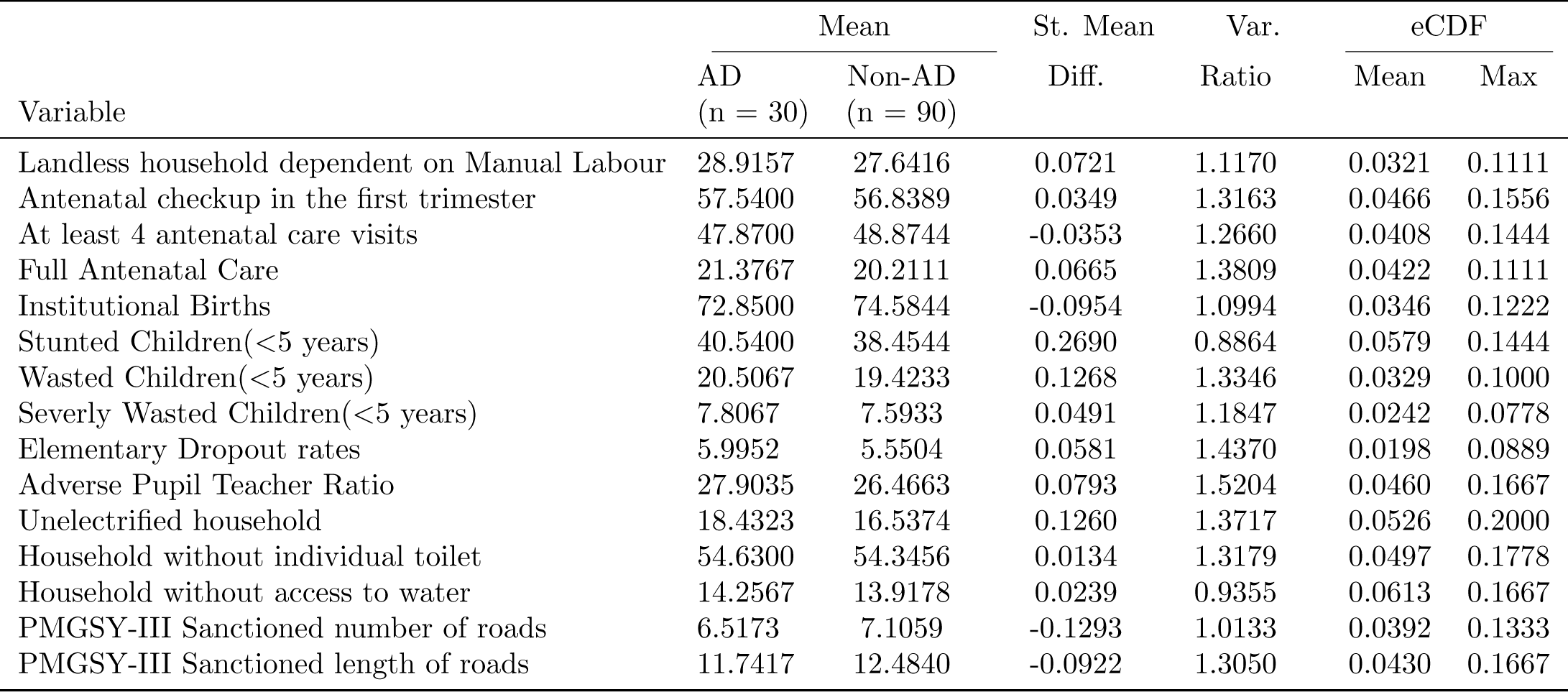
Summary of Covariate Balance after matching (Genetic Matching)

**Table S10:** Results of PSM-DID (Genetic Matching)

| No. | Indicators | Est. | St. err. | t-stat. | p-val. | Adj.<br>p-val. |
| --- | --- | --- | --- | --- | --- | --- |
| 1.3 | Registered pregnancies for which the mother received a Mother and Child Protection (MCP) card | 0.48 | 1.02 | 0.47 | 0.64 | 0.97 |
| 2 | Mothers who consumed iron folic acid for 100 days or more when they were pregnant | 3.43 | 3.18 | 1.08 | 0.28 | 0.97 |
| 4.1 | Sex ratio at birth for children born in the last five years | -3.16 | 32.10 | -0.10 | 0.92 | 0.97 |
| 5 | Home births that were conducted by skilled health personnel | 0.05 | 0.60 | 0.09 | 0.93 | 0.97 |
| 6.1 | Children under age 3 years breastfed within one hour of birth | 0.11 | 2.70 | 0.04 | 0.97 | 0.97 |
| 7 | Children under 5 years who are underweight (weight for age) | -3.05 | 0.96 | -3.17 | 0.00 | 0.02 |
| 8.2 | Children with diarrhoea in the 2 weeks preceding the survey who received oral rehydration salts (ORS) | -1.18 | 8.29 | -0.14 | 0.89 | 0.97 |
| 8.3 | Children with diarrhoea in the 2 weeks preceding the survey who received zinc | 1.75 | 5.40 | 0.32 | 0.75 | 0.97 |
| 8.4 | Children with fever or symptoms of ARI in the 2 weeks preceding the survey taken to a health facility or health provide | -0.60 | 3.46 | -0.17 | 0.86 | 0.97 |
| 10.1 | Breastfeeding children age 6-23 months receiving an adequate diet | 0.51 | 1.68 | 0.31 | 0.76 | 0.97 |
| 10.2 | Non-breastfeeding children age 6-23 months receiving an adequate diet | 4.78 | 5.75 | 0.83 | 0.41 | 0.97 |
| 11 | Children age 12-23 months fully vaccinated based on information from either vaccination card or mother's recall | 1.06 | 3.05 | 0.35 | 0.73 | 0.97 |
| 11 | Children age 12-23 months fully vaccinated based on information from vaccination card only | -0.92 | 3.96 | -0.23 | 0.82 | 0.97 |

## Footnotes

1 The mapping of the list of interventions under key programs to the targeted health indicators under ADP have been described in details in the table under the section Implementation for Improving Indicators on pages 44-48 of the Operational Guidelines^39^

2 The definitions of NFHS outcome variables that we include in our analysis is provided in table S2 in the appendix.

3 https://pib.gov.in/PressReleasePage.aspx?PRID=1845369

4 https://pib.gov.in/Pressreleaseshare.aspx?PRID=1680702

5 Pre-covid or NFHS-5 Phase 1 data was collected between 17 June 2019 to 30 January 2020 https://rchiips.org/nfhs/NFHS-5_FCTS/India.pdf

